# A transcriptomics-based meta-analysis combined with machine learning approach identifies a secretory biomarker panel for diagnosis of pancreatic adenocarcinoma

**DOI:** 10.1101/2020.04.16.20061515

**Authors:** Indu Khatri, Manoj K. Bhasin

## Abstract

Pancreatic ductal adenocarcinoma (PDAC) is largely incurable due to late diagnosis and absence of markers that are concordant with expression in several sample sources (i.e. tissue, blood, plasma) and platform (i.e. Microarray, sequencing). We optimized meta-analysis of 19 PDAC (tissue and blood) transcriptome studies from multiple platforms. The key biomarkers for PDAC diagnosis with secretory potential were identified and validated in different cohorts. Machine learning approach i.e. support vector machine supported by leave-one-out cross-validation was used to build and test the classifier. We identified a 9-gene panel (IFI27, ITGB5, CTSD, EFNA4, GGH, PLBD1, HTATIP2, IL1R2, CTSA) that achieved ∼0.92 average sensitivity and ∼0.90 specificity in discriminating PDAC from non-tumor samples in five training-sets on cross-validation. This classifier accurately discriminated PDAC from chronic-pancreatitis (AUC=0.95), early stages of progression (Stage I and II (AUC=0.82), IPMA and IPMN (AUC=1), IPMC (AUC=0.81)). The 9-gene marker outperformed the previously known markers in blood studies particularly (AUC=0.84). The discrimination of PDAC from early precursor lesions in non-malignant tissue (AUC>0.81) and peripheral blood (AUC>0.80) may facilitate early blood-diagnosis and risk stratification upon validation in prospective clinical-trials. Furthermore, the validation of these markers in proteomics and single-cell transcriptomics studies suggest their prognostic role in the diagnosis of PDAC.

## Introduction

Pancreatic ductal adenocarcinoma (PDAC) is the most common type of pancreatic cancer (PC), which is one of the fatal cancers in the world with 5-year survival rate of <5% due to the lack of early diagnosis (1). One of the challenges associated with early diagnosis is distinguishing PDAC from other non-malignant benign gastrointestinal diseases such as chronic pancreatitis due to the histopathological and imaging limitations (2). Although imaging techniques such as endoscopic ultrasound and FDG-PET have improved the sensitivity of PDAC detection but have failed to distinguish PC from focal mass-forming pancreatitis in >50% cases. Dismal prognosis of PC yields from asymptomatic early stages, speedy metastatic progression, lack of effective treatment protocols, early loco regional recurrence, and absence of clinically useful biomarker(s) that can detect pancreatic cancer in its precursor form(s) (3). Studies have indicated a promising 70% 5-year survival for cases where incidental detections happened for stage I pancreatic tumors that were still confined to pancreas (4, 5). Therefore, it only seems rational to aggressively screen for early detection of PDAC. Carbohydrate antigen 19-9 (CA 19-9) is the most common and the only FDA approved blood based biomarker for diagnosis, prognosis, and management of PC but it has several limitations such as poor specificity, lack of expression in the Lewis negative phenotype, and higher false-positive elevation in the presence of obstructive jaundice (3). A large number of carbohydrate antigens, cytokeratin, glycoprotein, and Mucinic markers and hepatocarcinoma– intestine–pancreas protein, and pancreatic cancer-associated protein markers have been discovered as a putative biomarkers for management of PC (6). However, none of these have demonstrated superiority to CA19-9 in the validation cohorts. Previously, our group discovered a novel five-genes-based tissue biomarker for the diagnosis of PDAC using innovative meta-analysis approach on multiple transcriptome studies. This biomarker panel could distinguish PDAC from healthy controls with 94% sensitivity and 89% specificity and was also able to distinguish PDAC from chronic pancreatitis, other cancers, and non-tumor from PDAC precursors at tissue level (7). The relevance of tissue-based diagnostic markers remains unclear owing to the limitations of obtaining biopsy samples. Additionally, most current studies are based on small sample sizes with limited power to identify robust biomarkers. Provided the erratic nature of PC, the major unmet requirement is to have reliable blood-based biomarkers for early diagnosis of PDAC.

The urgent need for improved PDAC diagnosis has driven a large number of genome level studies defining the molecular landscape of PDAC to identify early diagnosis biomarkers and potential therapeutic targets. Despite many genomics studies, we do not have a reliable biomarker that is able to surpass the sensitivity and specificity of CA19-9. The inherent statistical limitations of the applied approaches combined with batch effects, variable techniques and platforms, and varying analytic methods result in the lack of concordance (8). The published gene signatures of individual microarray studies are not concordant with comparative analysis and meta-analysis studies when standard approaches are used due to variability in analytical strategies (8).

In our work, we have included all the available gene expression datasets for PDAC versus healthy subjects from gene expression omnibus (GEO) (https://www.ncbi.nlm.nih.gov/geo/) and ArrayExpress database (https://www.ebi.ac.uk/arrayexpress/) measured via microarray or sequencing platforms. We have included the datasets derived from blood and tissue sources excluding cell lines in our analysis. The cell lines were excluded for they do not depict normal cell morphology and do not maintain markers and functions seen *in vivo*.

The approach of combining multiple studies has previously been stated to increase the reproducibility and sensitivity revealing biological insight not evident in the original datasets (9). Using the uniform pre-processing, normalization, batch correction approaches in the meta-analysis can assist in eliminating false positive results. Therefore, we used multiple datasets in combinations and further divided them in training, testing and validation sets to identify and validate the markers with secretory signal peptides. We hypothesize that proteins with secretory potential will be secreted out of the tissue into the blood and these markers can be used as prognostic markers in a non-invasive manner. There were no previous studies on identification of marker genes that could be used with least-invasive methods. Also, a set of multiple genes targeting different pathways and biological processes are more reliable and sensitive than single gene-based marker for complex diseases like cancer (8). We also corroborated the protein expression of our markers in proteomics datasets obtained from Human Protein Atlas (HPA) (https://www.proteinatlas.org/).

## Methods

### Dataset identification

The literature and the publicly available microarray repositories (ArrayExpress (https://www.ebi.ac.uk/arrayexpress/) and GEO (https://www.ncbi.nlm.nih.gov/geo/)) were searched for gene expression studies of human pancreatic specimens. The selected datasets were divided into five training sets and fourteen independent validation sets for initial development and validation of Biomarkers. To avoid the representation of the datasets only from tissues the few blood studies available were divided across all training and validation phase of this study.

Each training dataset (GSE18670, E-MEXP-950, GSE32676, GSE74629 and GSE49641) included a minimum of four samples of normal pancreas and a minimum of four samples of PDAC. In training set we included minimum two datasets with source pancreatic tissue and peripheral blood. This was done to identify a predictor based on genes that are detectable in both pancreatic tissue and blood. Datasets GSE18670 (Set1: 6 normal, 5 PDAC), GSE32676 (Set6: 6 normal, 24 PDAC) and E-MEXP-950 (Set3: 10 normal, 12 PDAC) was derived from pancreatic tissue, whereas GSE74629 (Set4: 14 normal, 32 PDAC) and GSE49641 (Set5: 18 normal, 18 PDAC) contain transcriptome profile of peripheral blood PDAC patients.

Further, 14 validation sets were also divided into three groups, one “Test sets” (**Table 1A**) and second “Validation Sets” (**Table 1A**) and third “Prospective Validation Sets” (**Table 1B**). Five Tissue studies were included: one from microdissected tissue samples (Set6: 6 normal, 6 PDAC) and four from whole tissues (Set7: 45 normal, 40 PDAC; Set8: 6 normal, 6 PDAC; Set9: 8 normal and 12 PDAC and Set10: 15 normal, 33 PDAC). One blood study from peripheral blood was also validated using the biomarker (E-Set11: 14 normal, 12 PDAC).

**Table 1A:** Datasets used for development and validation of secretory genes based PDAC classifier.

| Groups | Dataset | Normal | Tumor | Sample Type | Platform | Accession |
| --- | --- | --- | --- | --- | --- | --- |
| Training Sets | Set 1 | 6 | 5 | Enriched | U133 Plus 2.0 | E-GEOD-18670 |
|  | Set 2 | 6 | 24 | Whole Tissue | U133 Plus 2.0 | E-GEOD-32676 |
|  | Set 3 | 10 | 12 | Microdissected | U133A | E-MEXP-950 |
|  | Set 4 | 14 | 32 | Peripheral Blood | HumanHT-12 V4.0 | GSE74629 |
|  | Set 5 | 18 | 18 | Peripheral Blood | Gene St 1.0 | GSE49641 |
| Test sets | Set 6 | 6 | 6 | Microdissected | U133A | E-MEXP-1121 |
|  | Set 7 | 45 | 40 | Whole Tissue | Gene St 1.0 | GSE28735 |
|  | Set 8 | 6 | 6 | Whole Tissue | Gene St 1.0 | GSE41368 |
|  | Set 9 | 8 | 12 | Whole Tissue | U133 Plus 2.0 | E-GEOD-71989 |
|  | Set 10 | 15 | 33 | Whole Tissue | U133 Plus 2.0 | E-GEOD-16515 |
|  | Set 11 | 14 | 12 | Peripheral Blood | U133 Plus 2.0 | E-GEOD-15932 |
| Validation Sets | V1 | 61 | 69 | Whole Tissue | Gene St 1.0 | E-GEOD-62452 |
|  | V2 | 20 | 36 | Whole Tissue | U133 Plus 2.0 | E-GEOD-15471 |
|  | V3 | 9 | 45 | Whole Tissue | Agilent-028004 | GSE60979 |
|  | V4 | 12 | 118 | Whole Tissue | U219 | GSE62165 |
|  | V5 | 50 | 33 | Blood Platelet | HiSeq-2500 | GSE68086 |

**Table 1B:** Datasets used for prospective validation of secretory genes based PDAC classifier.

| Group | Dataset | Group | Pancreatic Tumor | Sample Type | Platform | Accession |
| --- | --- | --- | --- | --- | --- | --- |
| Prospective Validation Sets | PV1 | 4 Normal | 150 PDAC | Tissue | RNA-Seq | TCGA |
|  | PV2 | 61 Normal | 69 PDAC (Stage I and II) | Whole Tissue | Gene St 1.0 | E-GEOD-62452 |
|  | PV3 | 9 (Pancreatitis) | 9 (PDAC) | Whole Tissue | U95Av2 | E-EMBL-6 |
|  | PV4 | 7 (Normal) | 15 (IPMA, IPMC, IPMN) | Microdissected | U133 Plus 2.0 | GSE19650 |

For Phase I Validation we selected five datasets from different platforms from whole tissues and blood platelets, including comparison of normal versus PDAC samples similar to training and test sets. Four datasets from whole tissue (V1: 61 normal, 69 PDAC; V2: 20 normal, 36 PDAC; V3: 9 normal, 45 PDAC; and V4: 12 normal, 118 tumor) and one dataset from blood with samples from blood platelets (V5: 50 normal, 33 PDAC) were included.

In Prospective Validation, PDAC biomarker panel performance was tested on four additional independent datasets that compared results from: i) PDAC versus normal pancreatic tissue from TCGA database (PV1: 4 normal, 150 PDAC), ii) PDAC versus normal pancreatic tissues in early stages (PV2: 61 normal, 69 PDAC (Stage I and II)), iii) PDAC versus chronic pancreatitis (PV3: 9 pancreatitis, 9 PDAC), and iv) normal pancreas versus PDAC precursor lesions (intraductal papillary-mucinous adenoma (IPMA), intraductal papillary-mucinous carcinoma (IPMC) and intraductal papillary mucinous neoplasm (IPMN) with associated invasive carcinoma (PV4: 6 normal, 15 PDAC precursors (5 IPMA, 5 IPMC, 5 IPMN)) (**Table 1B**). Three datasets utilized oligonucleotide-based microarray platforms (two versions of Affymetrix GeneChips and Gene St 1.0 microarrays in one dataset) whereas TCGA data is sequencing data obtained using RNA-sequencing technology.

### Quality control and outlier analysis

Stringent quality control and outlier analysis was performed on all datasets used for training and validation to remove low quality arrays from the analysis. The technical quality of arrays was determined on the basis of background values, percent present calls and scaling factors using various bioconductor packages (10, 11). The arrays with high quality were subjected to outlier analysis using array intensity distribution, principal component analysis, array-to-array correlation and unsupervised clustering. The samples that were identified to be of low quality or identified as outliers were eliminated from the analysis.

### Mapping of platform specific identifiers to universal identifier

To facilitate the collation of the differentially expressed genes identified by analysis of individual datasets, the platform specific identifiers associated with each dataset were annotated to corresponding universal gene symbol identifiers. Gene Symbols were used in subsequent analyses including comparative analysis of different datasets as well as predictor development. Briefly Affymetrix data was annotated using the custom CDF from brainarray (http://brainarray.mbni.med.umich.edu). Affymetrix probe set IDs that could not be mapped to an Entrez Gene ID (GeneID) were removed from the gene lists. For Agilent-028004, HumanHT-12 V4.0 and Gene St 1.0 studies the raw matrix was directly retrieved from the GEO interactive web tool, GEO2R, which were further processed and normalized. The normalized and annotated genes for TCGA was obtained from Broad GDAC Firehose database (http://gdac.broadinstitute.org). We have removed 29 non-PDAC samples from tissue cancer genome atlas (TCGA) during validation as our classifier was trained using PDAC samples(12).

### Pre-processing and normalization of microarray datasets

Potential bias introduced by the range of methodologies used in the original microarray studies, including various experimental platforms and analytic methods, was controlled by applying a uniform normalization, preprocessing and statistical analysis strategy to each dataset. Raw Microarray dataset were normalized using vooma (13) algorithm which estimates the mean-variance relationship and use the relationship to compute appropriate gene expression level weights. Similarly, RNASEQ datasets were normalized using voom algorithm (14). The normalized datasets were used for performing meta-analysis as well as predictor development.

### Differential gene expression analysis for generating Meta-signature

To generate PDAC meta-signature, we performed differential expression analysis on individual datasets from training sets by comparing normal versus cancer samples. To identify differentially expressed genes, a linear model was implemented using the linear model microarray analysis software package (LIMMA) (15). LIMMA estimates the differences between normal and cancer samples by fitting a linear model and using an empirical Bayes method to moderate standard errors of the estimated log-fold changes for expression values from each probe set. In LIMMA, all genes were ranked by t statistic using a pooled variance, a technique particularly suited to small numbers of samples per phenotype. The differentially expressed probes were identified on the basis of absolute fold change and Benjamini and Hochberg corrected P value (16). The genes with multiple test corrected P value <0.05 were considered as differentially expressed. Comparative analyses were performed to identify those genes that are significantly differentially expressed across multiple PDAC datasets. Genes that are concordantly over or under expressed in three PDAC datasets (two tissues and one blood study) were included in PDAC meta-signature.

### Secretory Gene Set Identification

To identify a non-invasive predictor based on genes with secretory potential we selected genes that had signal peptide for secretory proteins and no transmembrane segments (noTM). The Biomart package in R with quering the gene symbols to SignalP database facilitated the analysis. The Ensembl Biomart database enables users to retrieve a vast diversity of annotation data for specific organisms. After loading the library, one can connect to either public BioMart databases (Ensembl, COSMIC, Uniprot, HGNC, Gramene, Wormbase and dbSNP mapped to Ensembl) or local installations of these. One set of functions can be used to annotate identifiers such as Affymetrix, RefSeq and Entrez-Gene, with information such as gene symbol, chromosomal coordinates, OMIM and Gene Ontology or vice-versa.

### Training and independent validation of PDAC classifier using support vector machine

The upregulated secretory genes differentially expressed from PDAC meta-signature was used for training of PDAC classifier. Classifier was generated by implementing the support vector machines (SVM) approach using Bioconductor and using 0 as the threshold. Polynomial kernel was used to develop all the models. SVM was first tuned using 10-fold cross-validation at different costs and the best cost and gamma functions were later used to perform classification. Classifiers were trained using normalized, preprocessed gene expression values. Performance of classifiers in the training sets was evaluated using internal leave-one-out cross-validation (LOOCV). The performance of classifiers was measured using threshold-dependent (e.g. sensitivity, specificity, accuracy) and threshold-independent receiver operating characteristic (ROC) analysis. In ROC analysis, the area under the curve (AUC) provides a single measure of overall prediction accuracy. We developed biomarker panels of five to ten genes to develop highly accurate biomarker panels. The biomarker panel with the highest performance in the training sets was chosen for assessment of predictive power in six independent test datasets using threshold-dependent and -independent measures *i*.*e*. AUC.

#### Survival analysis

To determine the association of key genes with survival in PC, we performed survival analysis using the TCGA database (https://cancergenome.nih.gov/). The survival analysis was performed on PDAC mRNA of 150 patients (excluding samples related to normal tissues and non-PDAC tissues (12)). Survival analysis was performed on the basis of individual mRNA expression using the Kaplan-Meier (K-M) approach (17). The normalized expression data for each gene was divided into high and low median groups. The survival analysis was performed using Kaplan-Meier analysis from survival package in R. The results of the survival analysis were visualized using K-M survival curves with log rank testing. The results were considered significant if the P values from the log rank test were below 0.05. The effects of mRNA on the event were calculated using univariate Cox proportional hazard model without any adjustments.

### Pathways analysis

The biological pathways for the genes was performed using ToppFun software of ToppGene suite (18). ToppGene is a one-stop portal for gene list enrichment analysis and candidate gene prioritization based on functional annotations and protein interactions network. ToppFun detects functional enrichment of the provided gene list based on transcriptome, proteome, regulome (TFBS and miRNA), ontologies (GO, Pathway), phenotype (human disease and mouse phenotype), pharmacome (Drug-Gene associations), literature co-citation, and other features. The biological pathways with FDR < 0.05 were considered significantly affected.

## Results

### PDAC Differential expression analysis and meta-signature development

To develop a gene based minimally-invasive biomarker for differentiating PDAC from normal/pancreatitis, we searched the publicly available databases GEO and ArrayExpress and literature mining. We identified 19 microarray and RNA sequencing studies containing PDAC and normal samples. These datasets were divided into training sets (for development of a PDAC biomarker classifier), independent test sets, validation sets and prospective validation sets (see overview of meta-analysis strategy in **Figure 1**). For classifier training, we performed meta-analysis on 3-tissue and 2-blood-based PDAC studies to identify meta-signature of genes that are consistently differentially expressed in blood and tissue during PC. To account for the differences in microarray/sequencing platform used in studies, we processed and normalised studies according to their platforms and the selected the genes that are common across various studies. The number of differentially expressed secretory genes ranged from 480 to 810 genes, totalling 2,010 significantly differentially expressed genes in the five training datasets. Venn diagram analysis of these differentially expressed genes identified 74 genes (35 downregulated and 39 upregulated) (**Table S1**) with concordant directionality to at least two of the three tissue datasets and one of the two blood datasets (**Figure 2A, shown in red color**).

**Figure 1:**
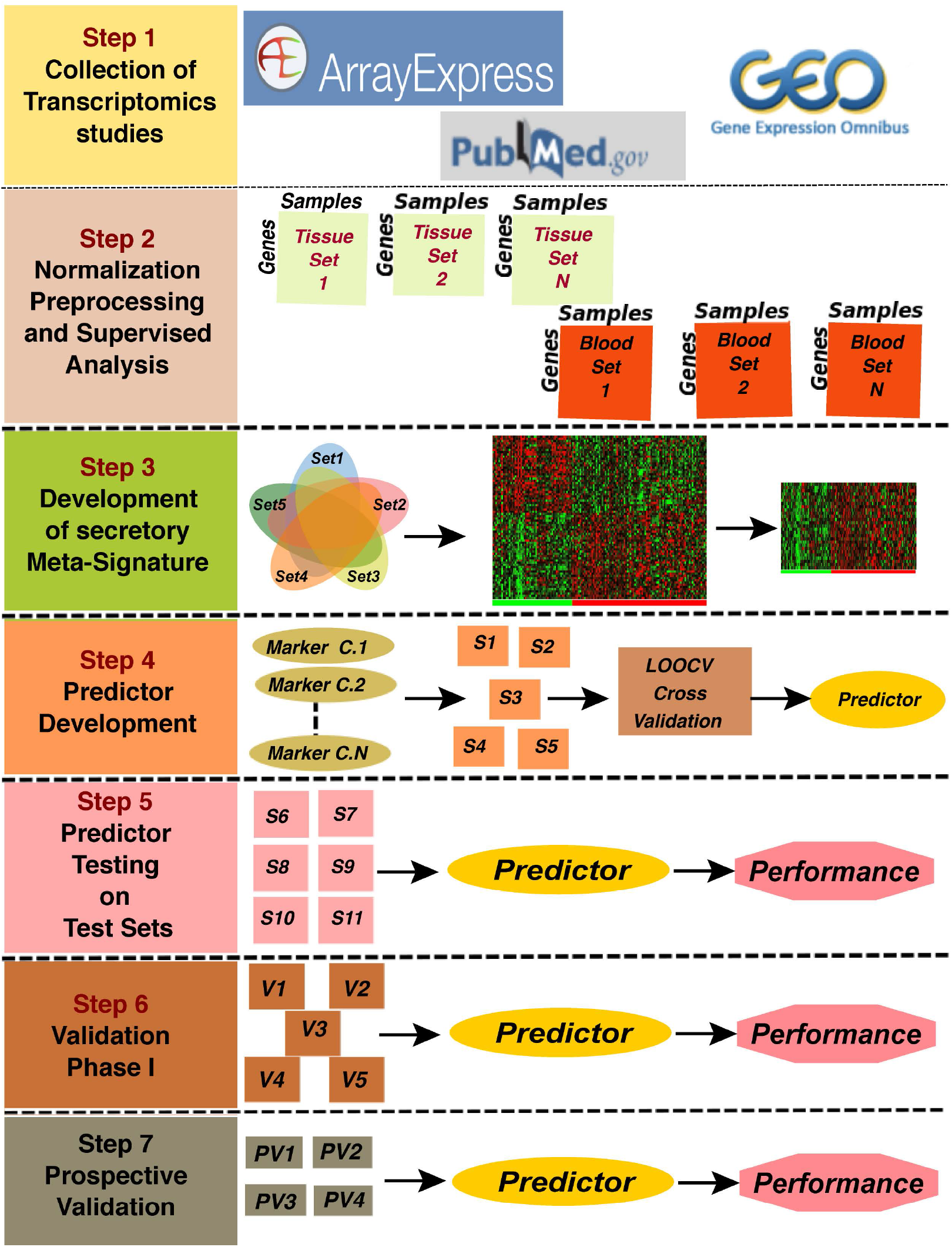
Overview of the meta-analysis approach for development and validation of PDAC biomarker panel. Predictor was developed using the data from Set1-Set5 (S1-S5 in Step 4) and was further tested on Set5-Set10 and validated on V1-V5 and PV1-PV4 datasets.

**Figure 2:**
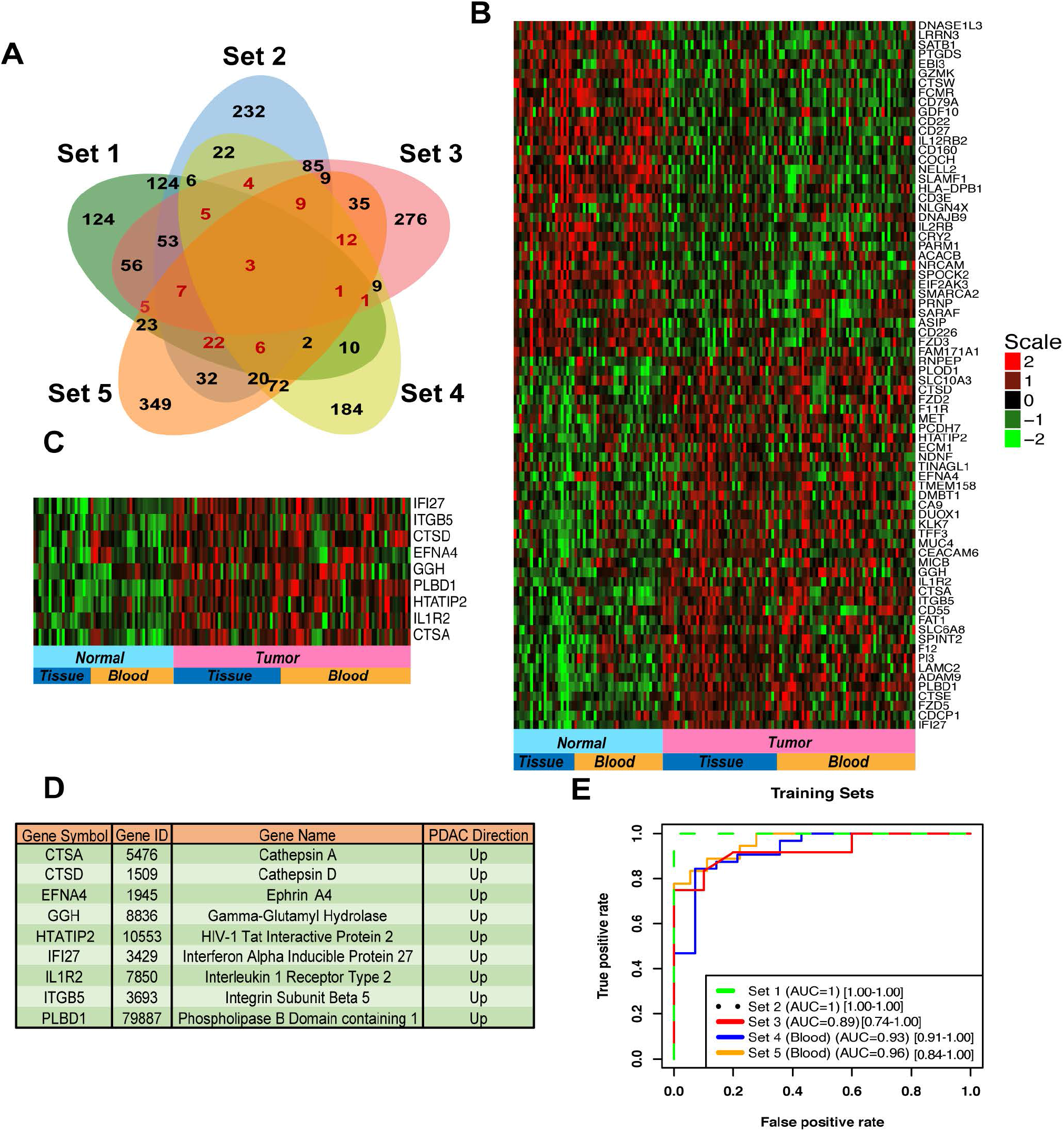
Meta-signature of genes that are consistently differentially expressed in multiple datasets and candidate PDAC diagnostic biomarker panel. **A**. Venn diagram of the five training datasets for the differentially expressed genes. 74 genes (marked in red) with concordant directionality are common to at least 2 of the 3 tissue datasets (Set 1 to Set 3) and one of the 2 blood datasets (Set 4 and Set 5). **B**. Heatmap of the 74 meta-signature genes differentially expressed in PDAC from five training datasets. Red = upregulated, Green = downregulated. **C**. Heatmap of the 9-upregulated marker genes in training sets for PDAC biomarker panel. **D**. Description of the genes from the 9-gene based PDAC biomarker panels. **E**. AUC plot [CI: 95%] for 9-gene PDAC classifier across the five training sets using leave one out cross-validation (LOOCV). Set1 and Set 2 are matched normal samples i.e. obtained from same individual. Set 3 normal samples are not matched, Normal samples are obtained from the patients undergoing surgery with other pancreatic diseases. Set 4 and Set 5 are blood sourced studies therefore the normal subjects were matched for gender, age and habits.

Consistent expression across these five datasets for each of the 74 concordant genes is demonstrated in a heatmap of the relative ratio of gene expression in PDAC compared to normal pancreas (**Figure 2B**), with the extent of over-expression or under-expression denoted by red or green shading, respectively. Pathway analysis of these 74 common PDAC genes depicted significant enrichment (P value <0.05) in multiple extracellular matrix associated pathways (e.g. Ensemble of genes encoding extracellular matrix and extracellular matrix-associated proteins, remodelling of the extracellular matrix, structural ECM glycoproteins, Cell adhesion molecules) (**Figure S1**). These pathways play important roles in the adhesion of cells that is a key process in progression of PDAC.

### Variables Selection and class prediction analysis in training sets

The 39-upregulated genes from the 74 common genes were selected for predictor development. We have specifically targeted upregulated genes for their therapeutics and diagnostic applications. We plotted boxplots of these 39 genes across all the five training sets and removed the genes with opposite direction in any of these five sets. The 27 concordantly upregulated genes (**Table S2**) were selected after the boxplot analysis. The heatmap for 27 genes (**Figure S2A**) and Principal Component Analysis (PCA) plots (**Figure S2B**) of these genes shows a separation pattern between PDAC and normal pancreas samples in each dataset. The predictors based on 5 to 10 genes were developed by implementing a SVM based classifier. Based on SVM with polynomial kernel and LOOCV evaluation in the training sets, classifiers containing 9 genes performed with highest accuracy (i.e., IFI27, ITGB5, CTSD, EFNA4, GGH, PLBD1, HTATIP2, IL1R2, and CTSA). These 9 genes across the five training sets demonstrate differential expression in PDAC compared to a normal pancreas across most of the samples (**Figure 2C, 2D**).

We performed LOOCV cross-validation analysis of the 9-gene PDAC classifier across the five training datasets to determine its predictive performance. For each of the five training datasets individually, sensitivity ranges from 0.83-1.0 and specificity 0.71-1.00 for the predictor (**Figure S3A, Table 2**). Comparison of the 9-gene PDAC classifier performance in tissues (Set1-Set3) and blood datasets (Set 4 and Set 5) shows an average 0.94 sensitivity and 0.97 specificity for the tissue datasets, in contrast to 0.88 sensitivity and 0.80 specificity for the blood datasets (**Figure S3B, Table 2**). AUC for the three tissue datasets ranged from 0.89-1.00 with median=0.96 **(Figure S3B)** and for two blood datasets from 0.92 to 0.96 with median=0.94 (Table 2, **Figure S3C and Fig 2E)**, demonstrates threshold independent performance). The average gene expression plots with all the samples combined from the five training sets (**Figure S4A**) and the PCA plots of training sets (**Figure S4B**) from 9 genes supports the discriminatory power of the marker combinations in identification of PDAC subjects from normal.

**Table 2:** The performance matrix of the 9-gene PDAC classifier on the training, testing, validation and prospective validation sets.

| Groups | Datasets | Accuracy | Sensitivity | Specificity | AUC |
| --- | --- | --- | --- | --- | --- |
| Training Sets | Set 1 | 1.00 | 1.00 | 1.00 | 1.00 |
|  | Set 2 | 1.00 | 1.00 | 1.00 | 1.00 |
|  | Set 3 | 0.87 | 0.83 | 0.90 | 0.89 |
|  | Set 4 | 0.82 | 0.93 | 0.71 | 0.93 |
|  | Set 5 | 0.86 | 0.83 | 0.89 | 0.97 |
| Test Sets | Set 6 | 1.00 | 1.00 | 1.00 | 1.00 |
|  | Set 7 | 0.92 | 0.90 | 0.93 | 0.94 |
|  | Set 8 | 1.00 | 1.00 | 1.00 | 1.00 |
|  | Set 9 | 0.95 | 0.91 | 1.00 | 1.00 |
|  | Set 10 | 0.96 | 0.93 | 1.00 | 0.94 |
|  | Set 11 | 0.73 | 0.75 | 0.71 | 0.80 |
| Validation Sets | V1 | 0.79 | 0.76 | 0.83 | 0.83 |
|  | V2 | 0.98 | 0.97 | 1.00 | 1.00 |
|  | **V3** | 0.94 | 1.00 | 0.89 | 0.98 |
|  | **V4** | 0.95 | 1.00 | 0.91 | 0.99 |
|  | **V5** | 0.83 | 0.84 | 0.82 | 0.89 |
| **Prospective Validation Sets** | **PV1** | 0.82 | 0.94 | 0.72 | 0.93 |
|  | **PV2** | 0.74 | 0.74 | 0.75 | 0.82 |
|  | **PV3** | 0.83 | 0.78 | 0.89 | 0.95 |
|  | **PV4-IPMA** | 1.00 | 1.00 | 1.00 | 1.00 |
|  | **PV4-IPMC** | 0.84 | 0.83 | 0.86 | 0.81 |
|  | **PV4-IPMN** | 1.00 | 1.00 | 1.00 | 1.00 |

### Significance of selected genes

CTSA and CTSD are involved in extracellular matrix associated proteins; IFI27 and IL1R2 in cytokine signalling in immune system; ITGB5 and HTATIP2 in apoptotic pathway and EFNA4, GGH and PLBD1 are involved in Ephrin signalling, fluoropyrimidine activity and glycerophospholipid biosynthesis respectively. The genes selected based on the presence of signal peptide for secretion are supposed to be secretory; however, the signal peptide is also present in several membrane proteins also (19). In the selected classifier genes, CTSD, EFNA4 and IL1R2 are predicted to be secretory proteins whereas CTSA, GGH, PLBD1, IFI27, ITGB5 and HTATIP2 are predicted to be intracellular or membrane bound proteins in HPA. Furthermore, CTSA and PLBD1 are also localized in Lysosomes and GGH is secretory protein as per UniProtKB (www.uniprot.org) predictions. Since our 9 gene markers could be detected with a detectable expression in both tissues and blood samples from PDAC patients, we further validated the performance of these genes for PDAC Diagnosis.

### Independent performance of classifier in differentiating PDAC from Normal

The biomarker set designed above was further tested in six independent sets with five tissue and one blood based PDAC studies. The classifier genes depicted an upregulation pattern in most of independent validation sets **Figure S5**. The boxplot revealed higher expression of all the 9 genes, averaged over test sets, in the tumor samples as compared to the healthy (**Figure 3A**). For each of the six datasets individually, sensitivity ranges from 0.75-1.00 and specificity from 0.71-1.00 for the predictor (**Figure 3B, Table 2**). Comparison of the 9-gene PDAC classifier performance in tissue and blood shows an average 0.94 sensitivity and 0.97 specificity for the tissue datasets, in contrast to 0.75 sensitivity and 0.71 specificity for the blood dataset. AUC for the five tissue datasets ranged from 0.94-1.00 and for one blood datasets AUC was 0.80 (**Figure 3C, Table 2)**.

**Figure 3:**
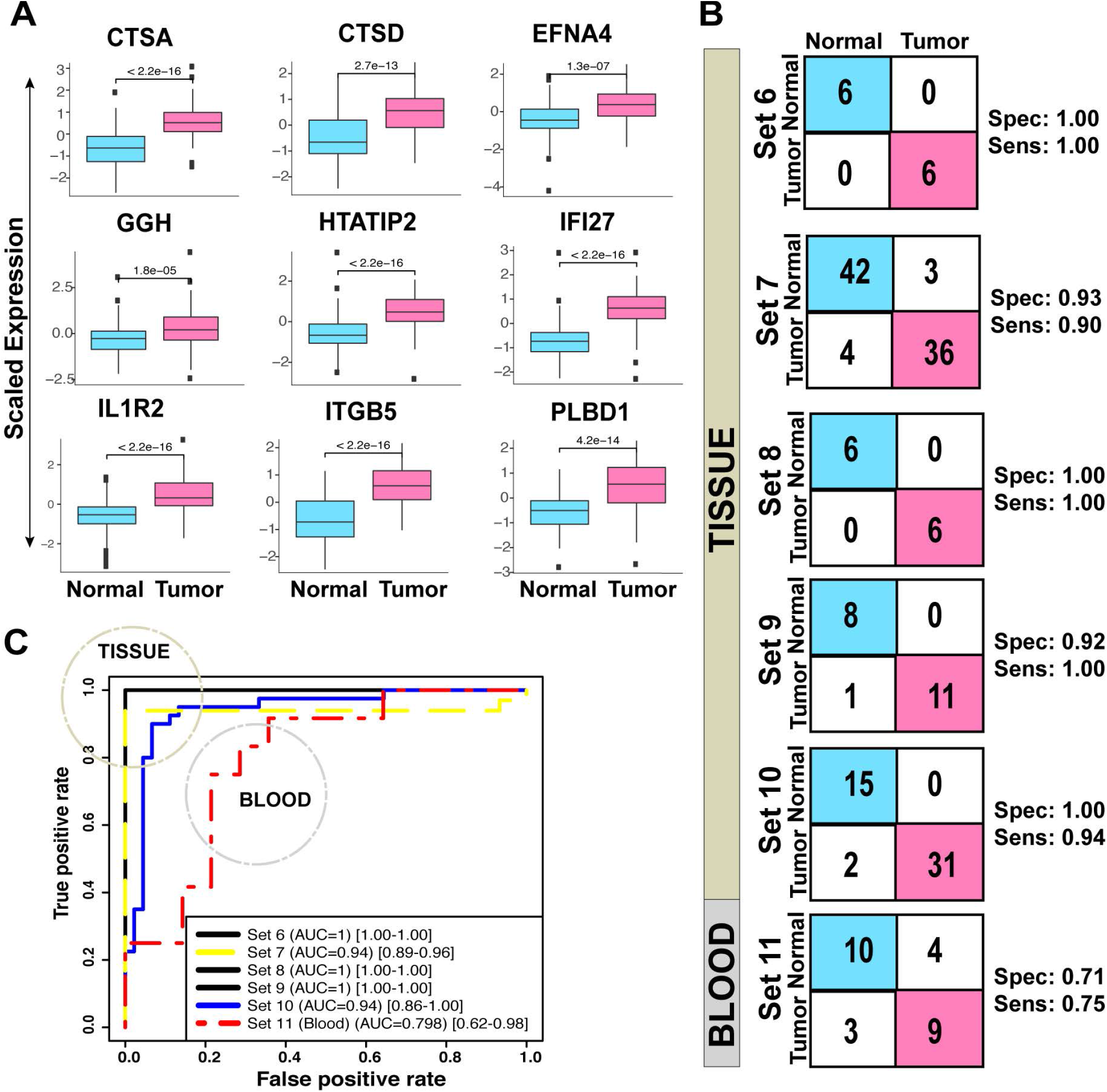
Performance of 9-gene PDAC Classifier on test sets using leave one out cross-validation (LOOCV). **A.** The boxplot of the averaged expression of the genes across all the six test datasets. The P values as calculated by t.test between the groups are on the individual genes. **B**. Diagnostic performance of the 9-gene PDAC classifier on the six test sets of PDAC vs. normal pancreas. Sensitivity (Sens.) and specificity (Spec.) indicated besides each set. **C**. AUC plot for 9- gene [CI: 0.95-0.99] PDAC classifier across the six test datasets.

### The 9-gene PDAC classifier predicts PDAC with high accuracy in 5 independent validation sets

In five validation sets, the 9-gene PDAC classifier accurately predicted the class of PDAC compared to normal with maximum AUC of 1.00 in the independent validation tissue (V2) set that contained 20 normal and 36 PDAC samples. More than 0.95 AUC was observed in three independent validation tissue sets (V2, V3 and V4) that contained 36, 45 and 118 PDAC and 20, 9 and 12 normal pancreas samples, respectively (**Figure 4A and Table 1B**). The boxplot revealed higher expression of all the 9 genes, averaged over validation sets, in the tumor samples as compared to the healthy samples (**Figure 4B**). In a tissue dataset (V1) containing 61 normal and 69 tumor samples a specificity of 0.83 and sensitivity of 0.76 was determined. In 50 normal and 33 PDAC blood platelet sample (V5) 0.84 sensitivity, 0.82 specificity and 0.88 AUC was achieved. The prediction of the PDAC class in comparison to normal was accurate with a sensitivity ranging 0.76-1.00 and specificity ranging between 0.82 and 1.00 (**Figure 4C panel II, Table 2**). **Figure S6** presents the heatmap of the nine genes in individual validation datasets and the PCA plots depicting the discrimination of PDAC from normal samples.

**Figure 4:**
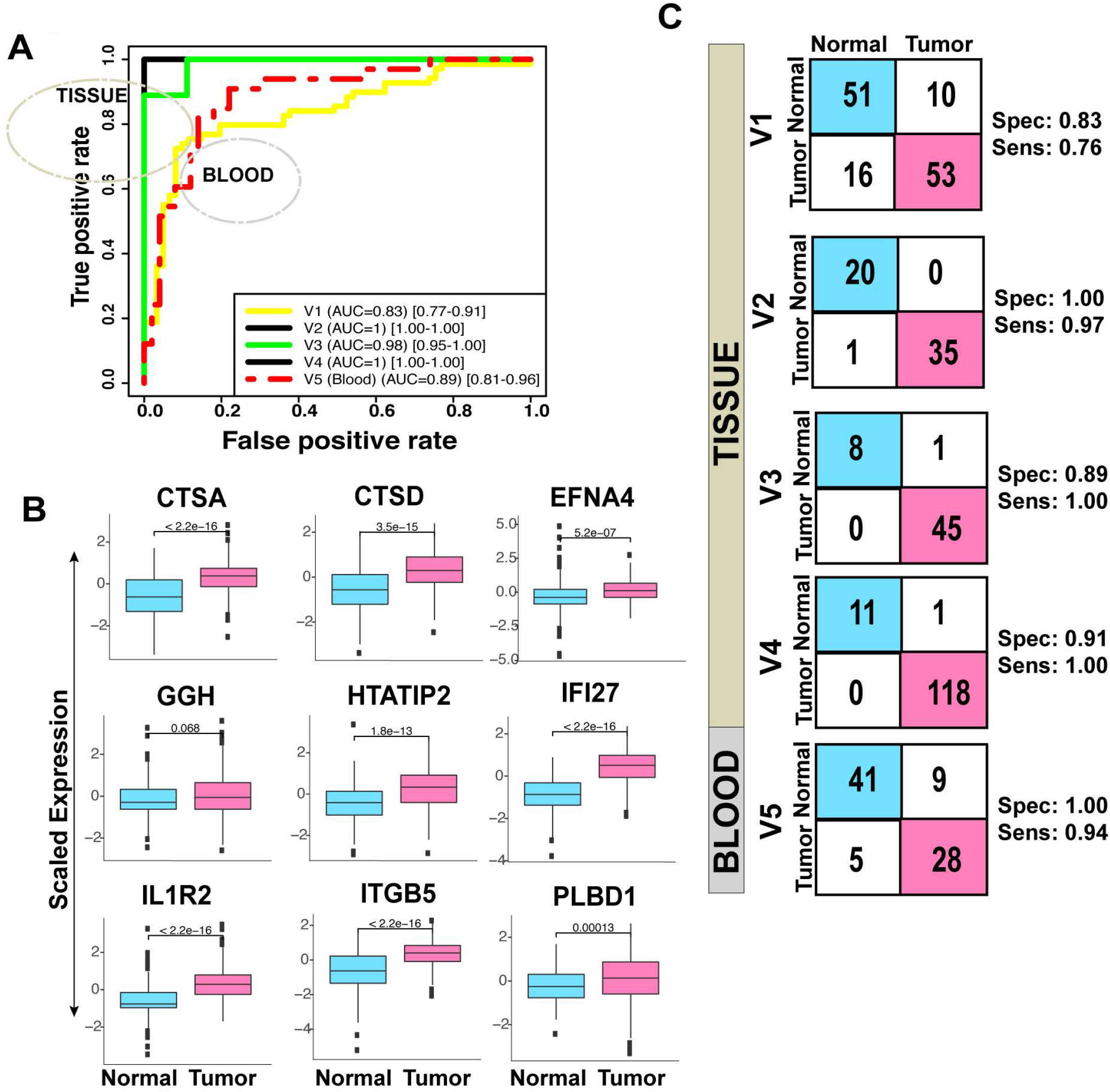
Performance of 9-gene PDAC Classifier on validation sets using leave one out cross-validation (LOOCV). **A**. The boxplot of the averaged expression of the genes across all the five validation datasets. The P values as calculated by t.test between the groups are mentioned on the individual genes. **B**. Diagnostic performance of the 9-gene PDAC classifier on the five validation sets of PDAC vs. normal pancreas. Sensitivity (Sens.) and specificity (Spec.) indicated besides each set. **C**. AUC plot [CI: 0.95-0.99] for 9-gene PDAC classifier across the five validation datasets.

### Cross-Platform Performance of Classifier on TCGA pancreatic samples

We further estimated the cross-platform performance of classifiers on the most widely used PC sample resource namely TCGA. TCGA datasets contain 150 PDAC samples and 4 normal samples and gene expression pattern analysis is not in consistence with other studies (**Figure S7C**). The cross-platform validation of classifier on TCGA data also achieved high sensitivity (0.94) and specificity (0.72) indicating the stability of the classifier in handling the cross-platform variation in absolute gene expression signal (**Figure 5 PV1**). The classifier achieved an excellent AUC of 0.93 (**Table 2**). The lower specificity of TCGA datasets might be due to the limited number of normal samples in the dataset. Heatmap of the 9 genes and PCA plots depicts the discrimination of two classes with the nine genes in the TCGA samples (**Figure S7 PV1)**.

**Figure 5:**
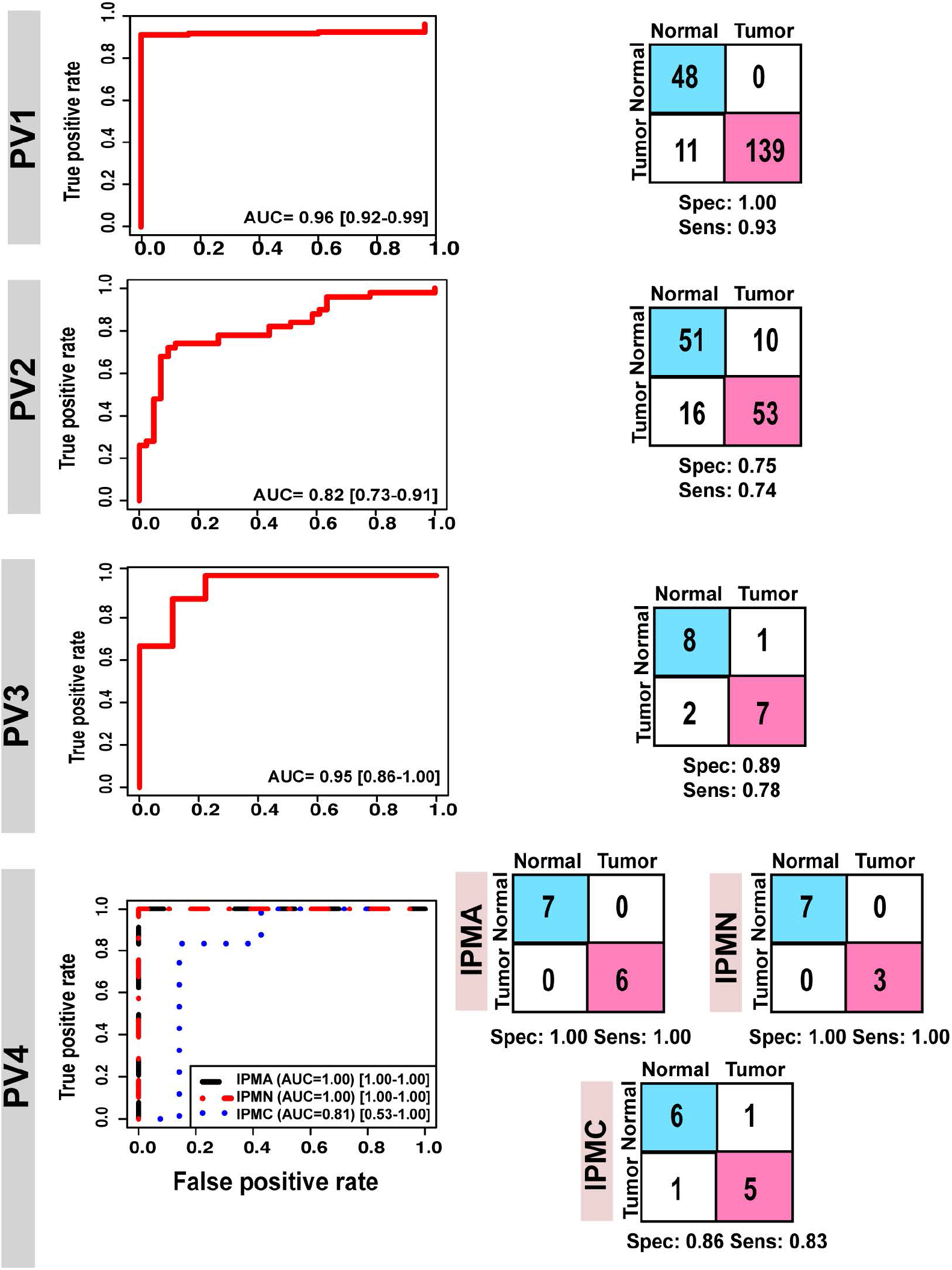
Performance of 9-gene PDAC Classifier on prospective validation sets using leave one out cross-validation (LOOCV). AUC plot [CI: 0.95-0.99] for 9-gene PDAC classifier and the diagnostic performance of **A**. the classifier for PV1 dataset, **B**. the classifier for PV2 dataset. **C**. the classifier for IPMA, IPMC and IPMN subjects in PV4 dataset and **D**. the classifier for PV3 dataset.

The markers did not show concordance in the TCGA dataset; however, the significance of these genes in the survival analysis can be very well established using the TCGA database. The samples were partitioned at median for selected nine-genes and survival analysis was performed on two clusters **(Figure S8)**. The results showed the combined survival of genes was able to clearly discriminate between better and poor survivors (P value significance of 0.05 and Hazard Ratio of 0.85), indicating their prognostic role in PDAC. High CTSD, EFNA4, HTATIP2, IFI27, ITGB5 and PLBD1 expression is associated with shortened survival time. Also, the survival analysis of these genes with a Hazard ratio of >1 at significant P value indicate their prognostic importance.

### Performance of Classifier in identifying early stage PDAC

As it is well established in literature that lack of established strategies for **early detection** of PDAC result in poor prognosis and mortality, we therefore tested performance of our classifiers on stage I and II PDAC. The predictor could distinguish stage I & II PDACs from normals with 0.74 sensitivity and 0.75 specificity and an AUC 0.82 (**Figure 5 PV2, Table 2**). Heatmap of the nine genes and PCA plots depicts the discrimination of two classes with the nine genes in early stages PDAC samples (**Figure S7 PV2)**.

### Performance of classifier in discriminating PDAC from Pancreatitis

Since discrimination between chronic pancreatitis (CP) and PDAC is a key clinical challenge, the fact that the 9-gene PDAC classifier accurately distinguishes between PDAC and CP is a further important validation step for this 9-gene biomarker panel. The array U95Av2 have the recorded signal intensity values for all the genes except PLBD1, hence only 8 genes were tested as a classifier for the discrimination of CP from PDAC. We tested the biomarker on the PV3 dataset wherein there were nine samples each for CP and PDAC. The classifier genes on PV3 dataset depicted significantly altered expression pattern between PDAC from CP (**Figure S7 PV3)**. The classifier achieved a specificity of 0.89 and sensitivity of 0.78 with an overall accuracy of 0.83 and an AUC of 0.95 in discriminating PDAC from CP (**Figure 5 PV3, Table 2**).

### Classifier discriminated pre-cancerous lesions from normal pancreas with good accuracy

To estimate the ability of the biomarker panel in discriminating precancerous lesions from a normal pancreas, we tested its performance on independent dataset containing laser microdissected normal main pancreatic duct epithelial cells and neoplastic epithelial cells from potential PDAC precursor lesions, IPMA, IPMC and IPMN [15]. Classifier genes were consistently overexpressed in the PDAC precursor samples, GGH was under-expressed in IPMA samples whereas it was overexpressed across the other PDAC precursors, IPMC and IPMN (**Figure S9**). The 9-gene PDAC classifier separates all potential PDAC precursor (IPMA, IPMC, IPMN) samples from the normal pancreatic duct samples except for one normal sample and one IPMC sample (**Figure 5 PV4**). The biomarker panel differed IPMA and IPMN from normal pancreatic duct epithelial cells with 1.00 sensitivity and 1.00 specificity, achieving an AUC of 1.00 (**Figure 5 PV4**). The predictor separated IPMC with 0.83 sensitivity and 0.86 specificity, achieving an AUC of 0.81 (**Table 2**).

### Classifier performed better than previous known markers

To estimate the performance of our current marker as compared to the previously established markers we compared the performance of our marker with each study [Bhasin et al (7), Balasenthil et al (20), Kisiel et al (21) and Immunovia (22)]. We used polynomial kernel for each set of markers and selected best model to record the performance on all the training, test and validation datasets (**Figure S10 and Table S3**). We found that all the methods performed well in tissue biopsies samples whereas when applied to the blood studies the performance of our marker set is the best (**Figure 6**). Our set of markers has performed well in tissues as well as blood studies and will be an ideal minimally invasive biomarker for studying in future studies and clinical trials.

**Figure 6:**
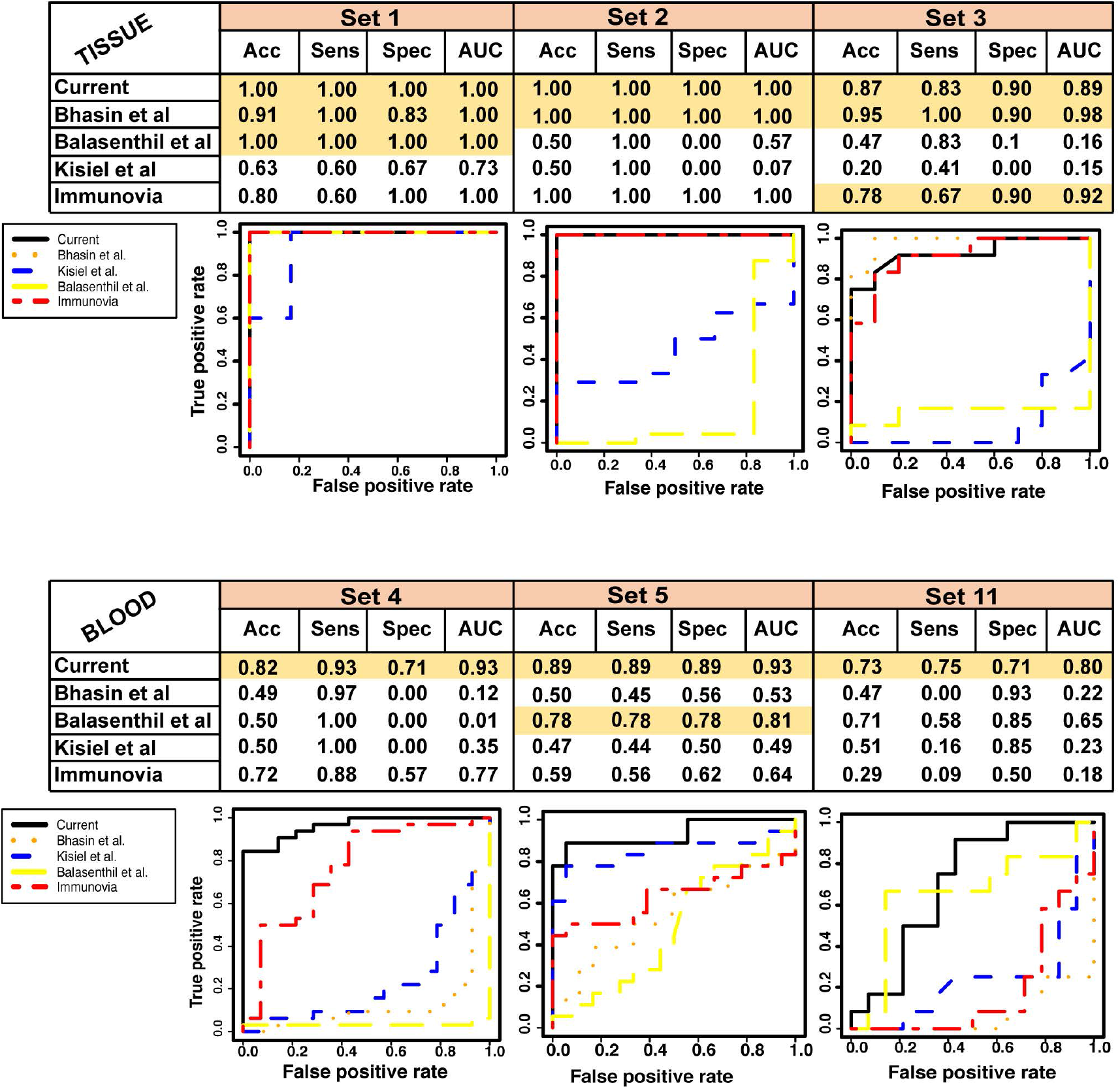
Comparative performance of 9-gene PDAC Classifier with different previously established biomarkers. AUC plot [CI: 0.95-0.99] for 9-gene PDAC classifier across the three tissue and three blood datasets. The boxes colored in mustard color have greater than 0.80 AUC.

### Validation of the markers in single-cell transcriptomics studies

Furthermore, as the markers are derived from bulk sequencing protocols it is important to know if the markers discovery is not influenced by different cell-types in normal and cancerous pancreas. Therefore, we used single-cell RNA-Seq data published by Peng et al (23) suggesting heterogeneity in PDAC tumor to plot expression of our markers on different cell-types. Using standard Seurat single-cell analysis methodology (24, 25), we identified that our markers are not associated with any cell-types and are expressed across major cell types in pancreatic cancer (**Figure S11)**. All our markers depicted upregulation in various tumor microenvironment cells including immune cells and endothelial cells.

### Validation of markers in blood-based proteomics study

The nine-gene markers in the classifier are discovered and validated from the transcriptomics studies, hence the validation of their expression at the protein level is necessary. Therefore, we confirmed the expression of the nine genes at the protein level in publicly available proteomics studies and HPA. The immunolabeling of the proteins of the respective genes in HPA (**Figure S12**) suggest higher staining of the proteins in tumors as compared to the normal samples except IFI27 where the expression of the protein cannot be detected. To further validate the protein expression of our markers we searched for the corresponding proteins in multiple pancreatic cancer proteomics studies (26–32). CTSD, a cathepsin family protein, and Ephrin and Interferon gamma family markers are found to be highly expressed in multiple proteomics studies (33–35).

## Discussion

We applied a data mining approach to a large number of publicly transcriptome datasets followed by class prediction analysis and validation in independent datasets to discover candidate PDAC biomarkers (36, 37), which were secretory in nature. We explored the secretome of the PDAC from the differential gene sets, for the first time, to investigate an accurate secretory/ non-invasive biomarker panel for the PDAC diagnosis. We report here a 9-gene PDAC classifier that differentiated PDAC as well as the precursor lesions from the normal with high accuracy. This 9-gene PDAC classifier was validated in 12 independent human datasets. The 9-gene PDAC classifier encodes proteins with secretory potential in pancreas and few other tissues.

The 9-gene PDAC classifier performed well across multiple microarray platforms from different laboratories, using either whole tissue, microdissected tissue or peripheral blood. While over 2500 candidate biomarkers have been associated with PDAC and some of these candidates are in various stages of evaluation, only CA19-9 is FDA-approved for PDAC (38–40). Nevertheless, CA19-9 does not provide an accuracy high enough for screening, particularly for early detection or risk assessment. Currently, no diagnostic or predictive gene or protein expression biomarkers that accurately discriminate between healthy patients, benign, premalignant and malignant disease have been extensively validated. The goal of this study was to identify a biomarker panel with greater sensitivity and specificity corroborating across different sources and platforms.

Differential diagnosis between PDAC and pancreatitis is critical, since patients with CP are at increased risk of PDAC development and pathological discrimination between PDAC and pancreatitis can be challenging for definitive diagnosis of PDAC. The 9-gene PDAC classifier accurately distinguishes premalignant and malignant pancreatic lesions such as pancreatic intraepithelial neoplasia (PanIN), IPMN with low- to intermediate grade dysplasia, IPMN with high-grade dysplasia and IPMN with associated invasive carcinoma from healthy pancreas. We discovered that all 9 genes are overexpressed already in PanIN, indicating that these 9 genes become dysregulated very early during PDAC development and could indeed assist in the early detection of PDAC. An early detection marker, one able to detect PDAC precursor lesions (IPMN, PanIN) with early malignant transformation or high risk for malignant transformation, would increase the likelihood of identifying patients with localized disease amendable to curative surgery. Better diagnosis of borderline and invasive IPMNs and MCNs would be highly significant, and enable patients to choose the most appropriate course of action; this 9-gene PDAC classifier may provide such a risk assessment. Discovery and validation of a distinct set of sensitive and specific biomarkers for risk-stratifying patients at high risk for developing PDAC would eventually enable routine screening of high-risk groups (i.e., incidental detection of pancreatic lesions, family history of PDAC, hereditary syndromes, CP, type 3c diabetes, smokers, BRCA2 carriers, etc).

While other studies have performed meta-analysis of transcriptome data for PDAC to identify the genes that are overexpressed in PDAC (41–43), they are irrelevant in identifying the markers for prognosis of PDAC. A panel of five serum-based genes (44) highlighted the potential of including relevant mouse models to assist in biomarker discovery. On the other hand, there has been significant progress in identifying circulating miRNAs that distinguish PDAC from CP and healthy patients in plasma and bile [42]. A five-miRNA panel diagnosed PDAC with 0.95 sensitivity and specificity in a cohort that included healthy, CP and PDAC patients [42]. However, similar to gene studies, there is no evidence on whether these miRNAs would diagnose early stages of PDAC.

To determine whether the set of biomarkers encoded by our PDAC classifier may also reflect key pathophysiological pathways associated with PDAC development or progression that may be candidate therapeutic targets, we reviewed available public data for the classifier genes. Several genes of our 9-gene classifier have been linked to tumorigenesis, indicating a causal role in PDAC development and progression. HTATIP2 is involved in apoptosis function in liver metastasis related genes (45), gastric cancer (46) and pancreatic cancer (47). IFI27, functioning in immune system, has been suggested as a marker of epithelial proliferation and cancer (41, 48). ITGB5 involved in integrin signalling have been found to be upregulated in several analysis studies (49). The Integrin and ephrin pathways have been proposed to play an important role in pancreatic carcinogenesis and progression, including *ITGB1*, a paralog of *ITGB5*, and EPHA2 as most important regulators (49). EPHA2 belongs to ephrin receptor subfamily and is involved in developmental events, especially in the nervous system and in erythropoiesis. To this family belongs one of our genes EFNA4 which activates another ephrin receptor EPHA5. IL1R2 was identified as possible candidate gene in PDAC and as one of the two higher level defects of the apoptosis pathway in PDAC (50). Il1, the ligand of IL1R2 is secreted by pancreatic cells (51) and has important functions in inflammation and proliferation and can also trigger the apoptosis (52– 54). CTSD have been shown to be upregulated in the PDAC cancer (42). AGR2, a surface antigen, has been shown to promote the dissemination of pancreatic cancer cells through regulation of Cathepsins B and D genes (55). CTSA was identified as one of the 76 deregulated genes in a study aiming for the development of early diagnostic and surveillance markers as well as potential novel preventive or therapeutic targets for both familial and sporadic PDAC (56). PLBD1 has been found to be upregulated in various studies with five-fold increase in cell lines (57) and in study where the effect of pancreatic β-cells inducing immune-mediated diabetes was studies (58). Metabolism-related gene [γ-glutamyl hydrolase (GGH) has been found to relevant and upregulated in gallbladder carcinomas (59).

Most of the classifier genes (ITGB1, EPHA2, IL1R2) have been linked to migration, immune pathways, adhesion and metastasis of PDAC or other cancers, specifically associated with developmental events and signaling. However, these biological functions would be anticipated to be involved in PDAC progression and early stages of PDAC development. To corroborate this aspect in more detail we evaluated the expression levels of these “PDAC progression” genes in the transcriptome datasets comparing PDAC precursors (LIGD-IPMN, HGD-IPMN) and InvCa- IPMN to normal pancreas, and PDAC vs. PanIN vs. healthy pancreas in the GEM model) (**Figure 5**) [15]. Eight genes except GGH are overexpressed in LIGD-IPMN, HGD-IPMN, and InvCa- IPMN as well as in PanINs, as compared to a normal pancreas, demonstrating that enhanced expression of multiple genes linked to metastasis and PDAC progression occurs early on during malignant development. This analysis indicates that the PDAC classifier may reflect some driving early defects during PDAC development. This argument is further strengthened by the survival analysis of the genes where five of the nine genes (CTSA, CTSD, EFNA4, IFI27 and IL1R2) are strongly related to discriminating better and poor survivors.

Further, to analyse the potential of the 9-gene biomarker in accurate classification of PDAC subjects versus healthy subjects we compared our biomarker combination with previously known and established biomarker combinations. Our analysis also indicates that the multiplex panel of biomarkers, rather than a single biomarker, is more likely to improve the specificity and selectivity for accurate detection of PDAC. The idea behind generation of biomarker panel with the better identification in blood sample in corroboration with the tissue studies is fulfilled here. The previously established markers worked well in the tissue studies but could not show their similar potential in blood studies.

Further, the protein expression of selected biomarker genes was also examined to determine their association with PDAC at protein levels. The analysis depicted that multiple gene product/proteins corresponding to biomarkers genes depicted higher expression in pancreatic cancer tissues. Interestingly some marker (e.g., EFNA4, GGH) also depicted over-expression in other cancers indicating their association with tumor development and progression related hallmark processes. In recent years multiple proteomics studies were performed to understand the proteome landscape of the PDAC but still lack in generating comprehensive picture due to technological limitations. Most of the proteomics technique can measure the expression of 2,000-3,000 proteomics that is far from generating the global overview of proteome. High expression of Cathepsin family proteins specifically CTSD is noted in several proteomics studies which was also the case for Ephrin and Interferon gamma family markers (33–35). Also, the expression of these genes is not found to be related to a particular cell-type in pancreatic cancer cell lineage. However, the fact that the overall study is based on bulk sequencing data cannot be overlooked and these cells may comprise of multiple cell-types which may or may not influence the overall methodology of marker selection. Overall, the protein-expression of the selected genes and their expression in multiple cell-types of pancreatic cancer is established. However, the aforementioned limitations have to be challenged before designing the diagnostic panel.

The 9-gene markers identified here still needs validation in bigger cohort for its potential in identifying accurately the early stages but this marker combination potentially has shown its discriminatory power across various blood and tissue datasets obtained from different sources and different platforms.

## Data Availability

The datasets used and/or analysed during the current study are available in public repositories GEO and ArrayExpress. The codes and DE genes per dataset will be available via GitHub (https://github.com/IKhatri-Git/Secretory-gene-classifier).

## Abbreviations

AUC: area under the curve
CA 19-9: Carbohydrate antigen 19-9
CP: chronic pancreatitis
GEO: gene expression omnibus
GGH: γ-glutamyl hydrolase
HPA: Human Protein Atlas
IPMA: intraductal papillary-mucinous adenoma
IPMC: intraductal papillary-mucinous carcinoma
IPMN: intraductal papillary mucinous neoplasm
LOOCV: leave-one-out cross-validation
noTM: no transmembrane segments
PanIN: pancreatic intraepithelial neoplasia
PC: pancreatic cancer
PDAC: Pancreatic ductal adenocarcinoma
ROC: receiver operating characteristic
SVM: support vector machines
TCGA: tissue cancer genome atlas

## Declarations

### Ethical approval and Consent to participate

Not applicable

### Consent for publications

Not applicable

### Competing interests

BIDMC will be filling patent on behalf of MB and IK on the use of biomarker panel for early PDAC diagnosis. MB is an equity holder at BiomaRx and Canomiks.

### Funding

This study was supported through BIDMC CAO Innovation grant.

### Authors’ contributions

**IK** performed all the bioinformatics analysis and wrote the manuscript. **MB** supervised the bioinformatics analysis and edited the manuscript. Both the authors read and approved the final manuscript.

## Acknowledgements

Not applicable

## Supplementary Data

**Supplementary Figure S1.**
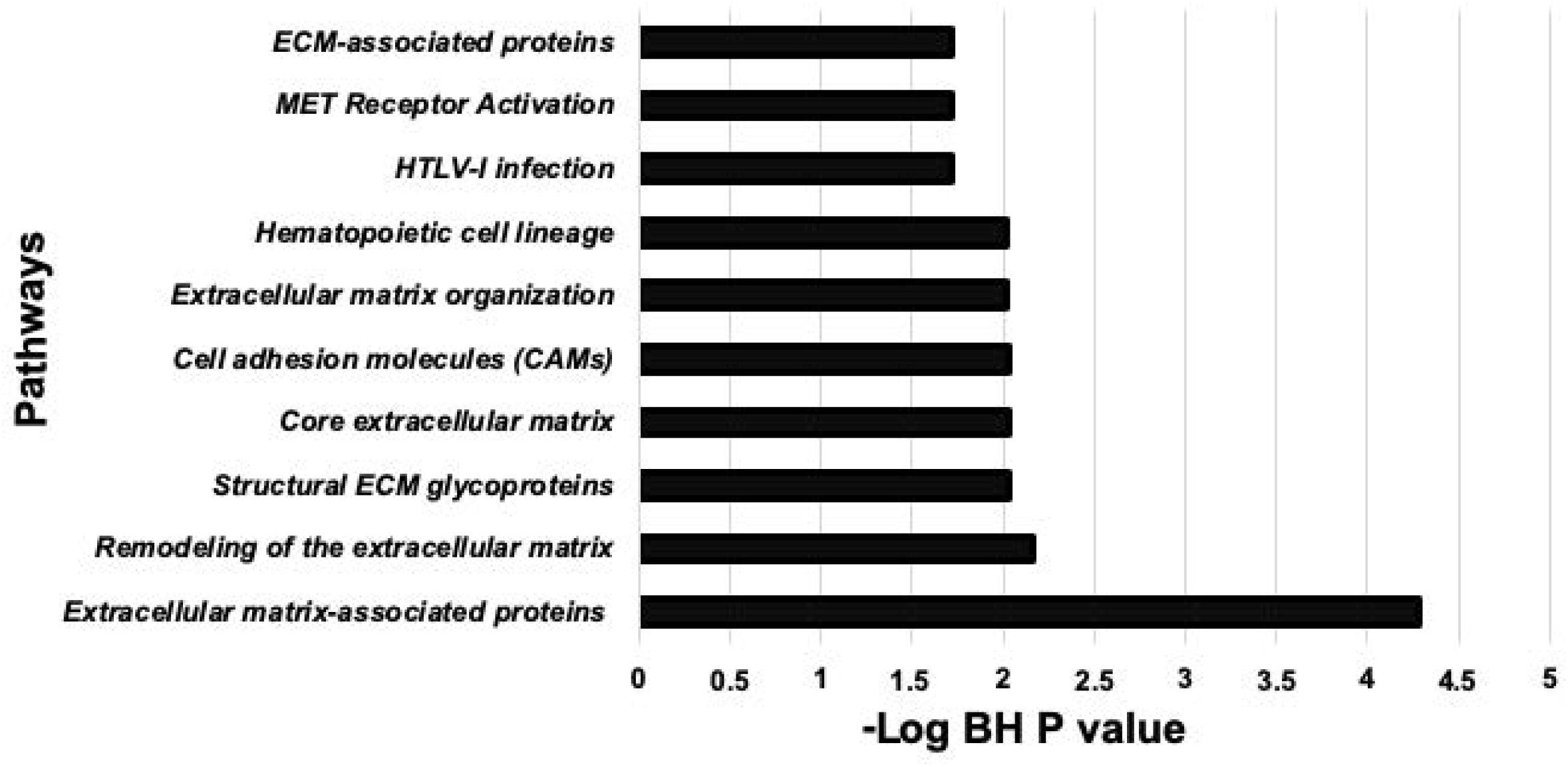
Pathway enrichment analysis of the 74 PDAC-specific secretory genes.

**Supplementary Figure S2:**
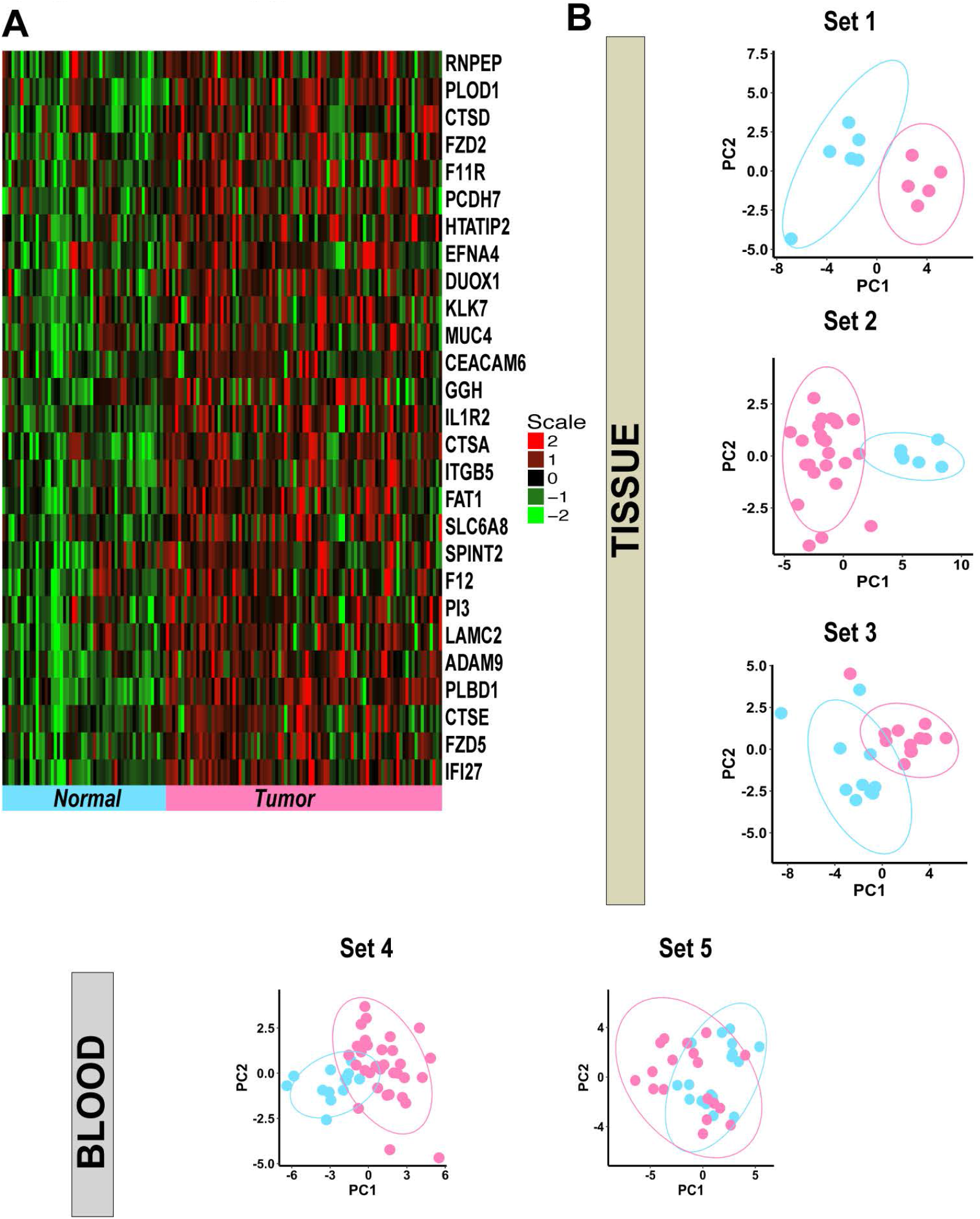
Upregulated Secretory genes in training datasets. **A)** Heatmap of 27 upregulated secretory genes in PDAC for two of the three tissues and one of the two blood datasets. **B)** PCA plots for each training datasets using 27 upregulated secretory genes.

**Supplementary Figure S3:**
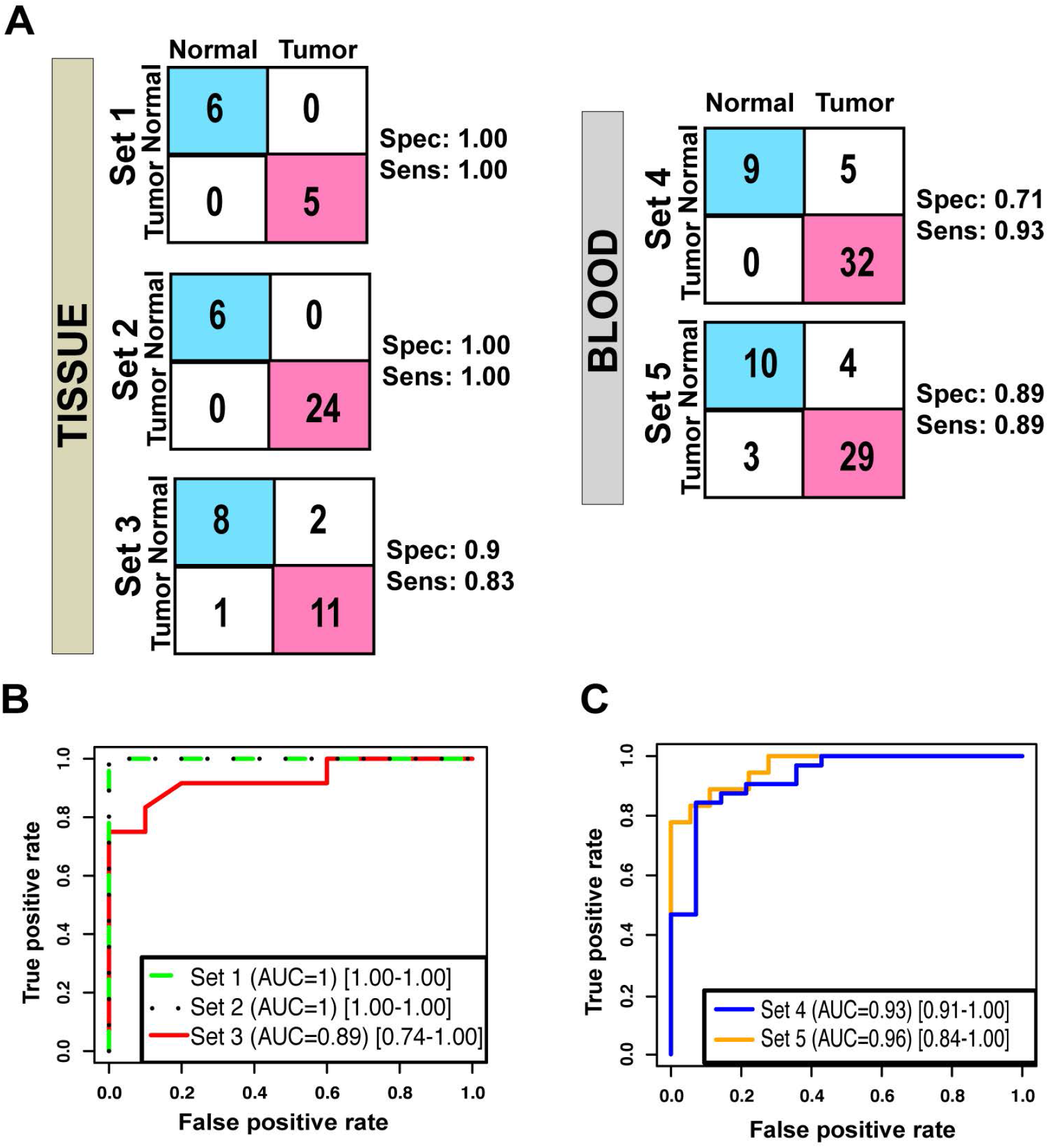
Performance of 9-gene PDAC classifier on training sets using leave one out cross-validation (LOOCV). **A)** Diagnostic performance of the 9-gene PDAC classifier on the five training sets. Sensitivity (Sens) and Specificity (Spec) are indicated for each dataset. B) AUC plot for 9-gene PDAC classifier on the three tissue training datasets. **C)** AUC plot for 9-gene PDAC classifier on the two blood training datasets.

**Supplementary Figure S4:**
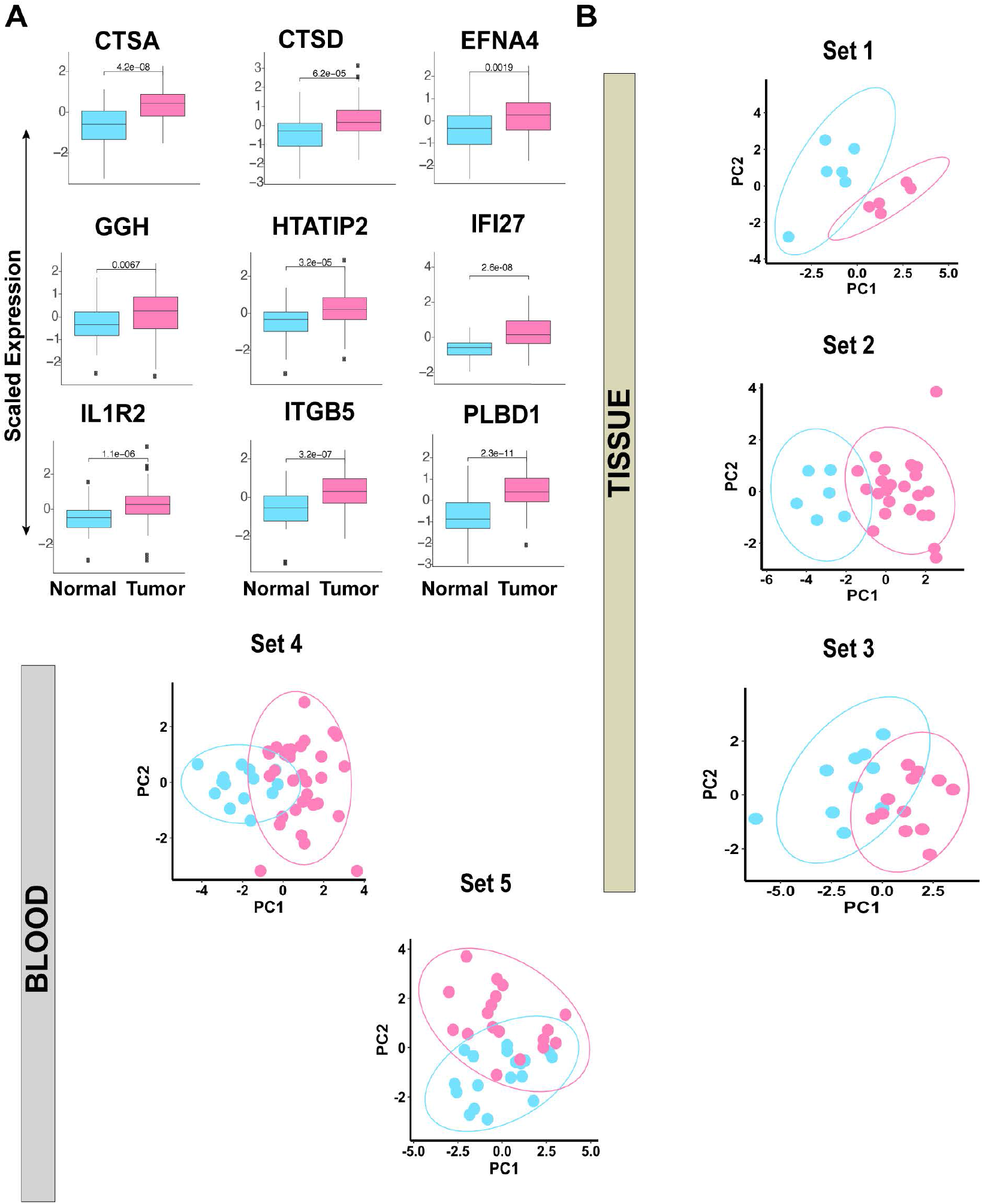
The metrics for training datasets using the 9-biomarker panel genes. **A)** Boxplot of the averaged expression of the genes across all the five training datasets. **B)** PCA plots for each training datasets using the 9-biomarker panel genes.

**Supplementary Figure S5:**
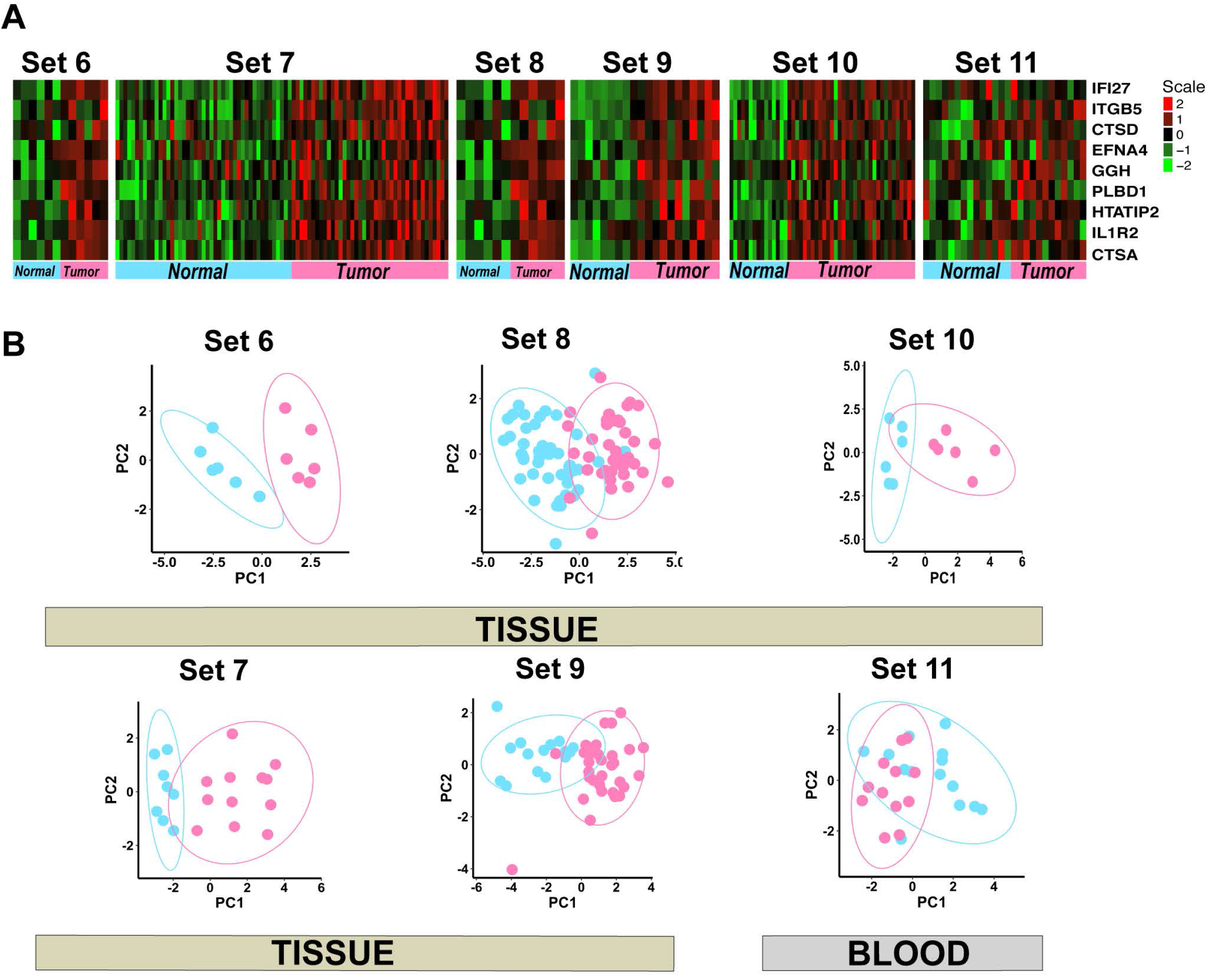
The assessment metrics for testing datasets using the 9-biomarker panel genes. **A)** Heatmap of the 9 PDAC-upregulated marker genes. **B)** PCA plots in six independent testing datasets.

**Supplementary Figure S6:**
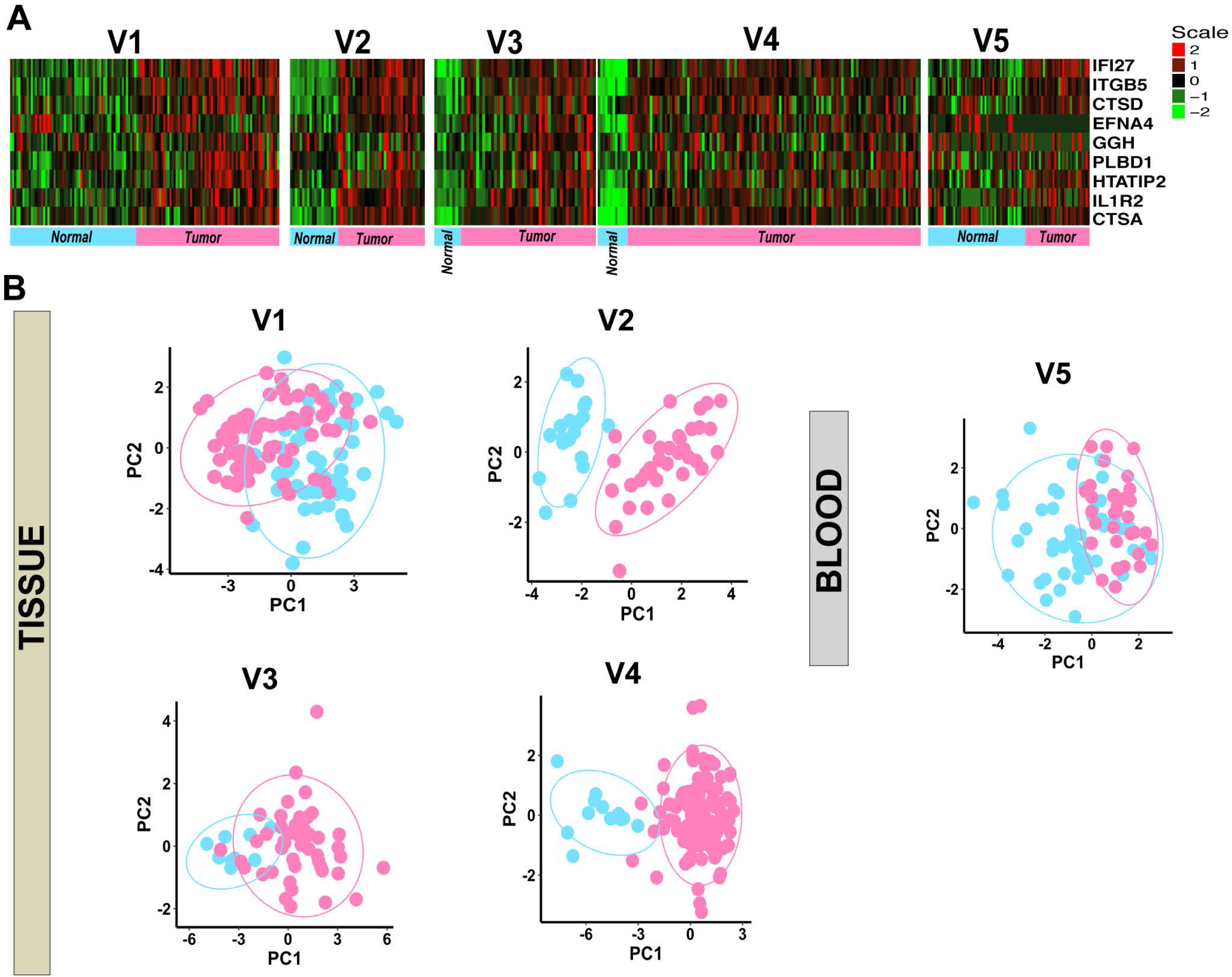
The assessment metrics for validation datasets using the 9-biomarker panel genes. Heatmaps **(A)** and PCA plots **(B)** based on biomarker panel genes in validation sets.

**Supplementary Figure S7:**
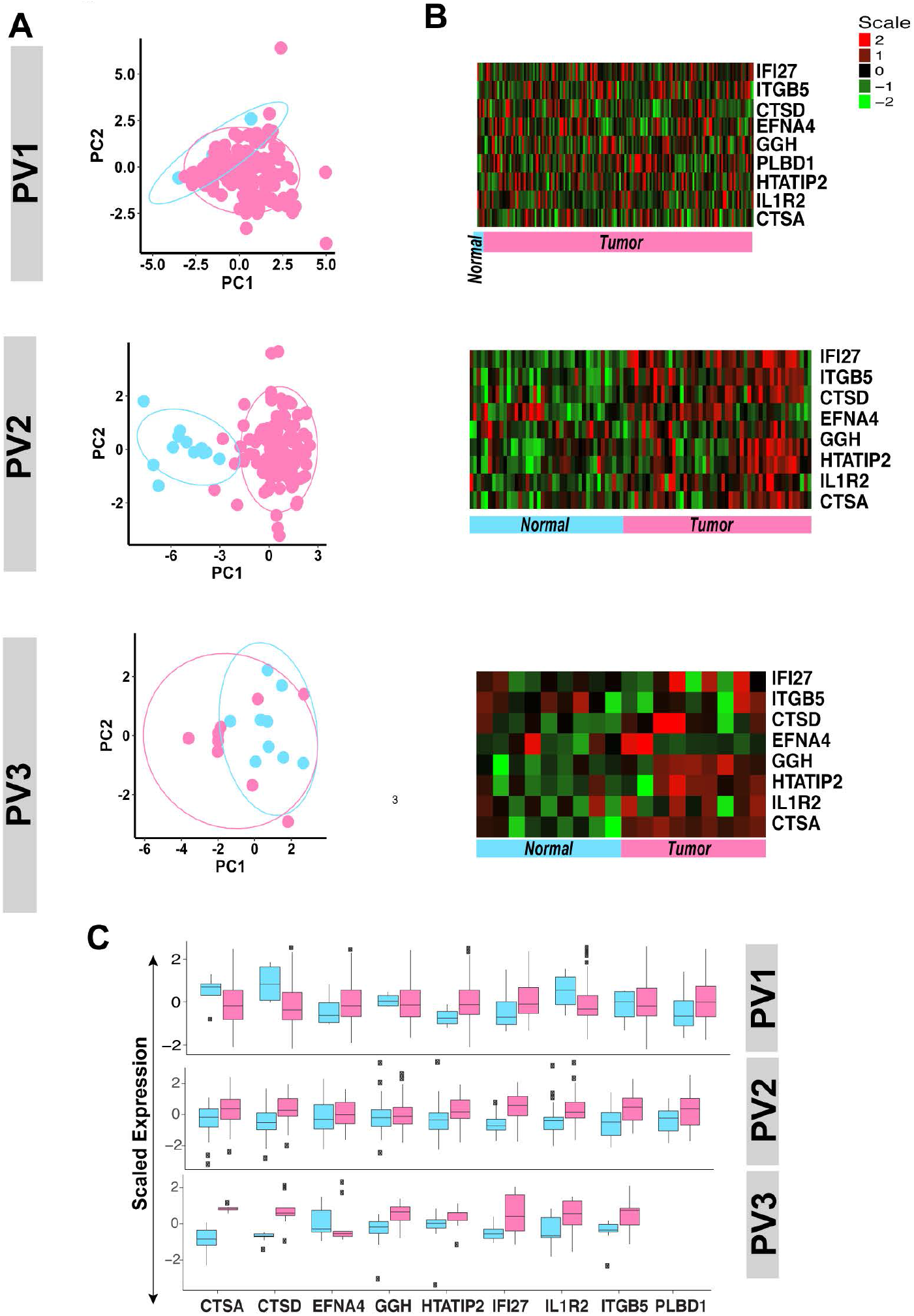
The assessment metrics for PV1-3 dataset using the 9-biomarker panel genes. **A)** PCA plots of three different prospective validation datasets. **B)** Heatmaps of the 9-marker genes panel. **C)** Boxplots of the expression of the genes.

**Supplementary Figure S8:**
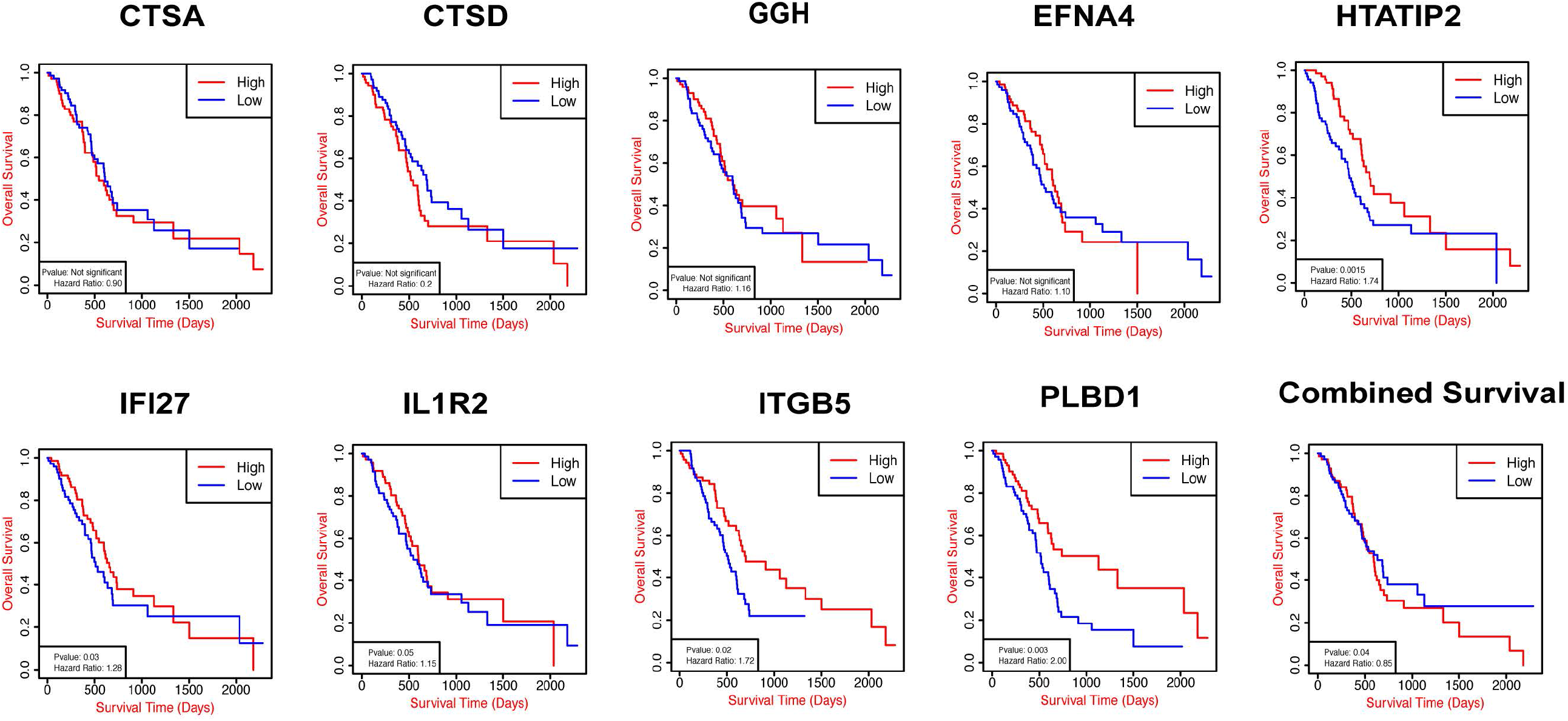
Survival curve of 9-gene-based PDAC classifier and combined genes.

**Supplementary Figure S9:**
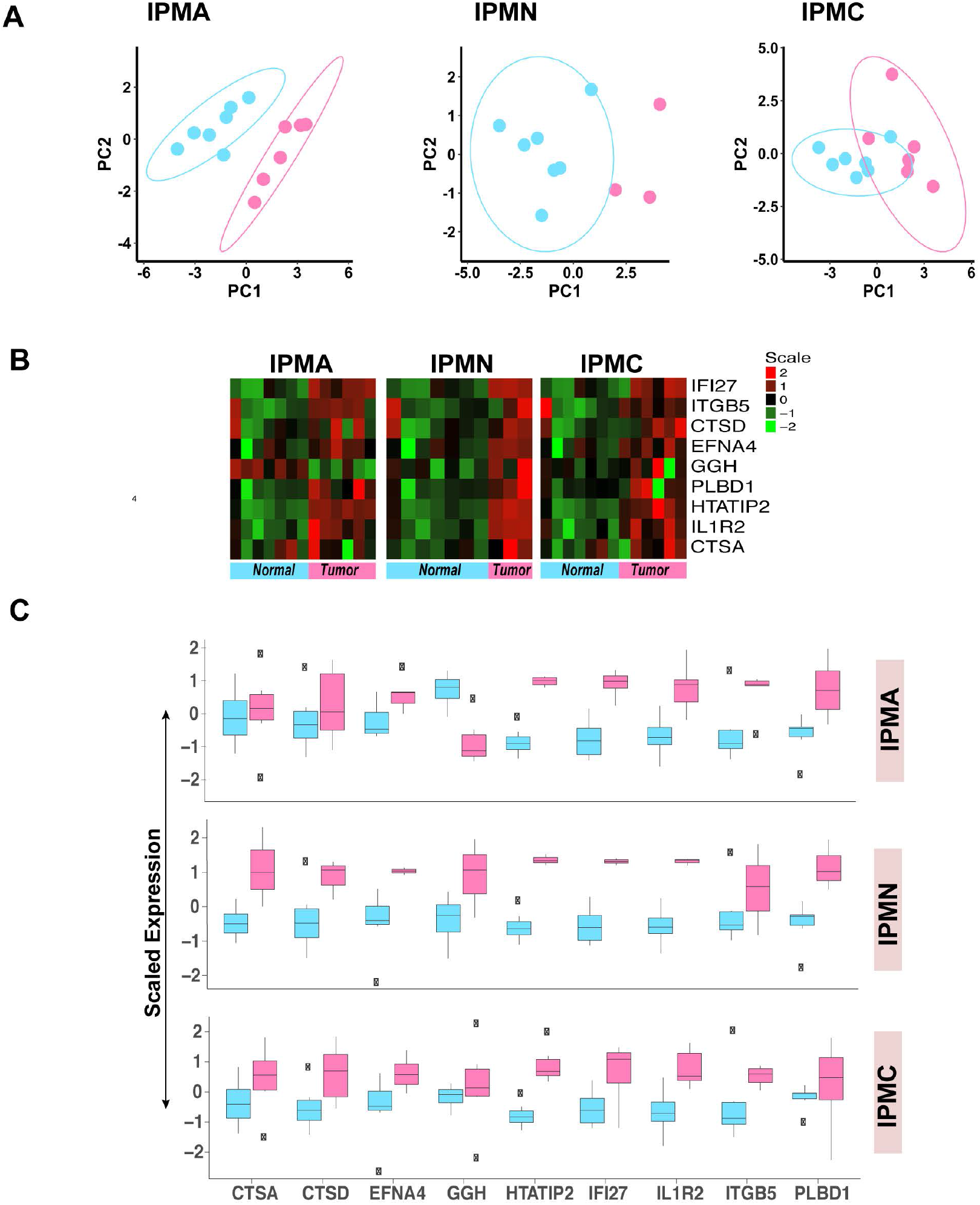
The assessment metrics for PV4 dataset using the 9-biomarker panel genes. **A)** PCA plots for precursor lesions in three stages IPMA, IPMN and IPMC. **B)** Heatmaps of the 9-marker genes panel. **C)** Boxplots of the expression of the genes in precursor lesions.

**Supplementary Figure S10:**
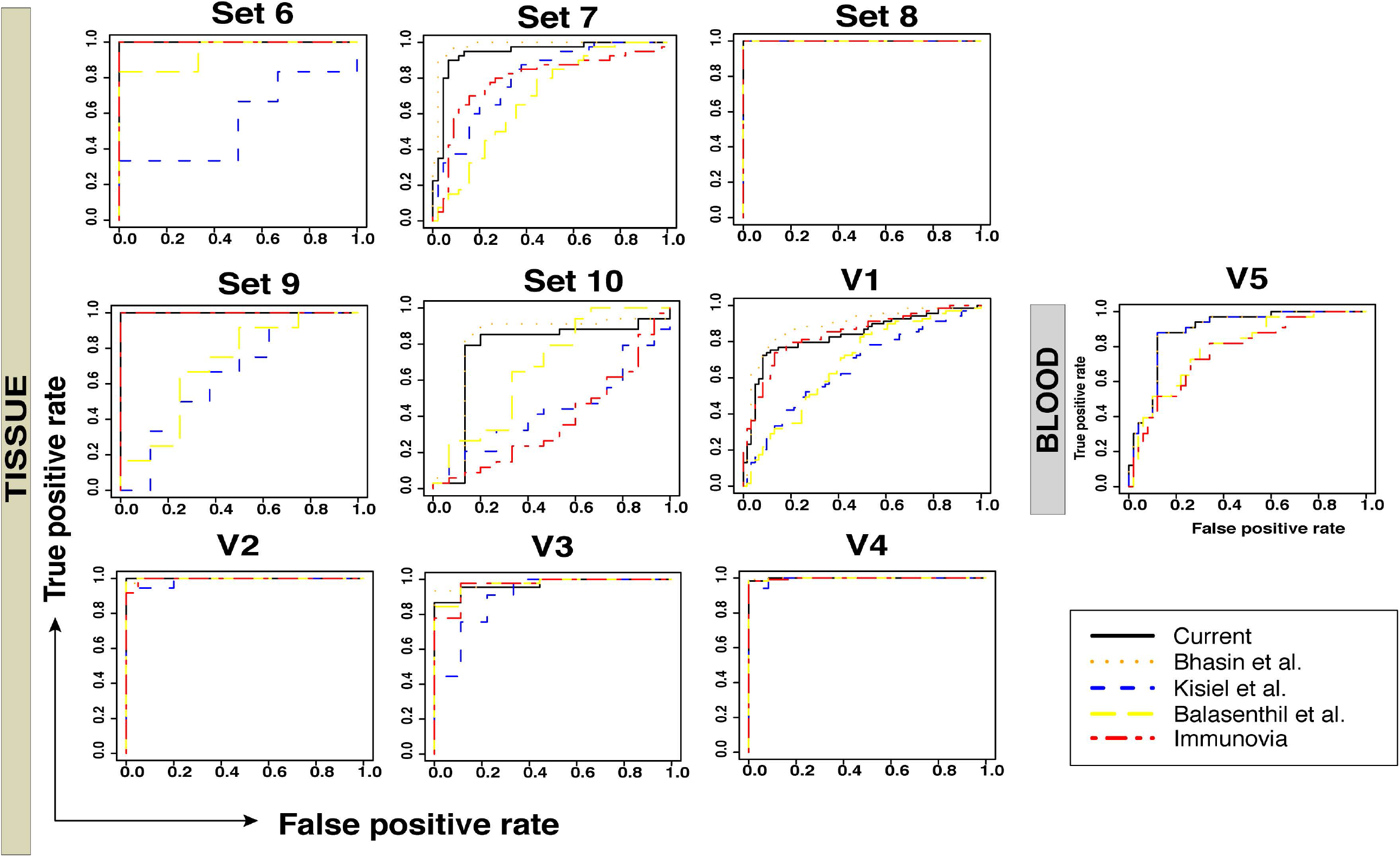
Comparative performance of 9-gene-based PDAC classifier with different previously established biomarkers. AUC plot for 9-gene-based PDAC classifier across the training and validation datasets. The measures of performances e.g. accuracy, sensitivity, specificity and AUC are mentioned in **Supplementary table 4**.

**Supplementary Figure S11:**
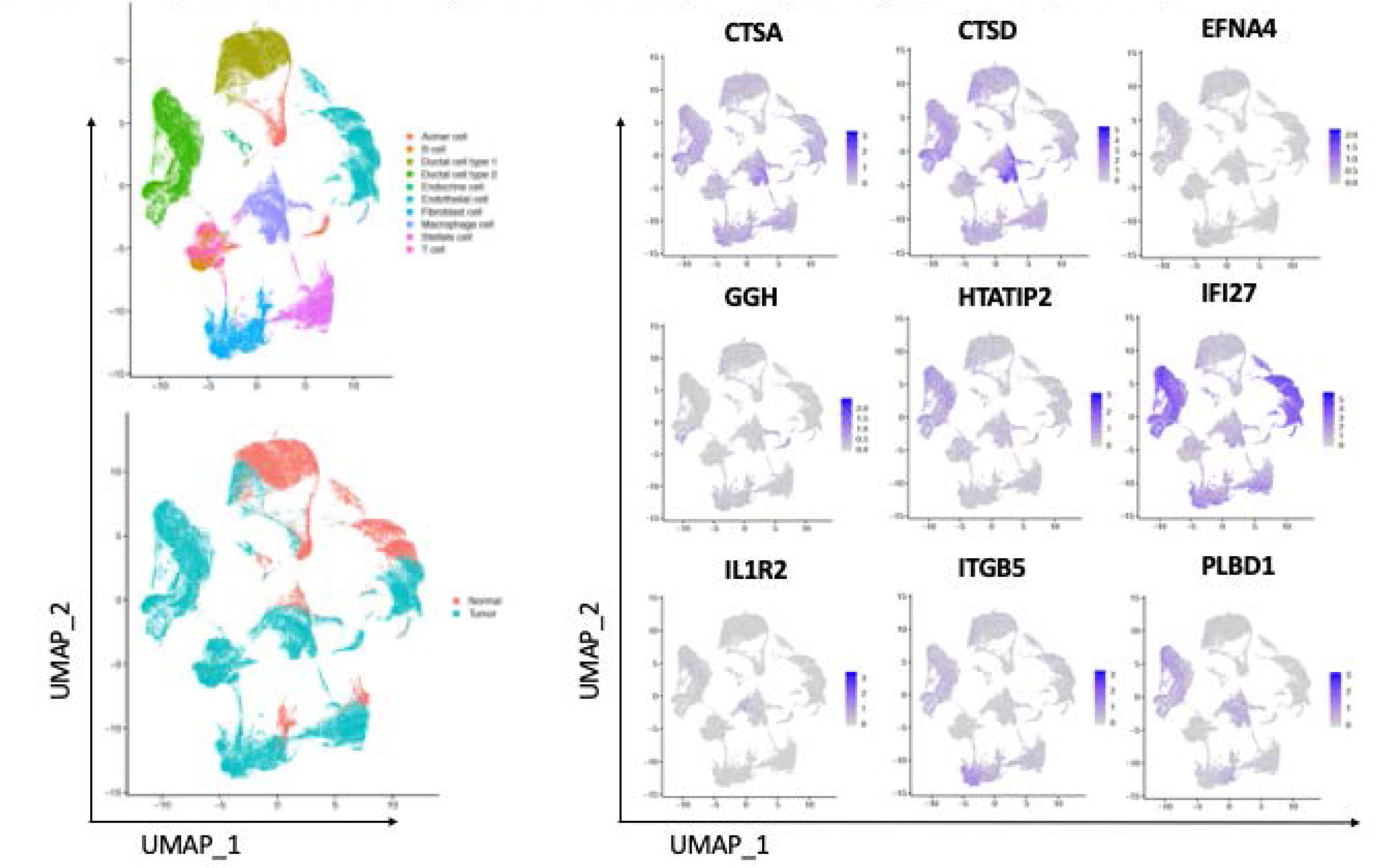
Expression of 9-gene markers in different pancreas cell-types in both healthy and tumor states. The expression of these genes is high in tumor state (CTSA, CTSD, EFNA4, GGH, HTATIP2, IFI27 and ITGB5) or they are not expressed at all in healthy state (IL1R2 and PLBD1) Source: Peng J et al., Cell Research, 2019^9^. This is also consistent with protein expression of the genes as measured by antibody staining experiments by Human protein atlas.

**Supplementary Figure S12:**
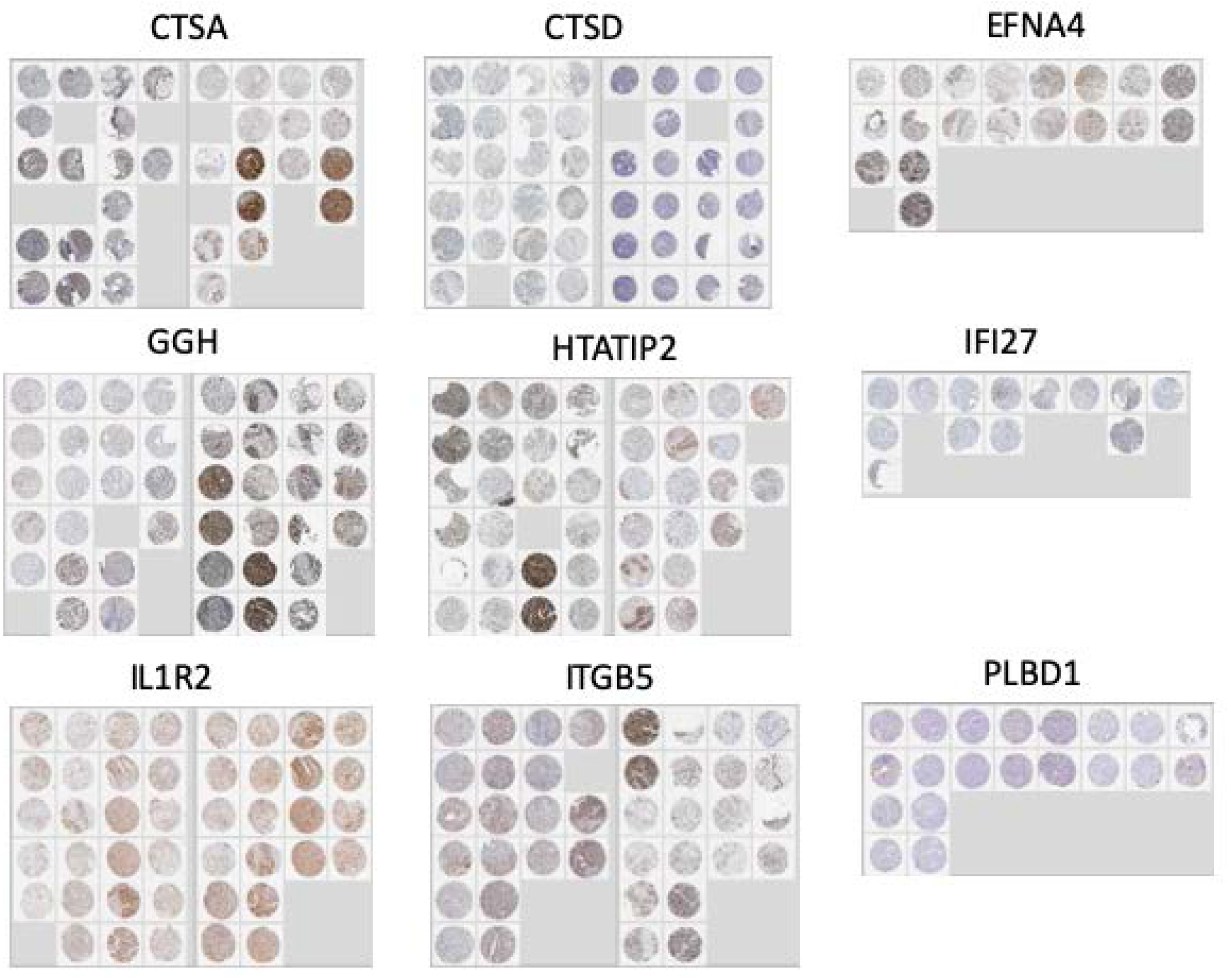
Immunolabeling of protein expression of nine genes selected for the classifier in pancreatic cancer. Light blue is low staining; blue is moderate staining and brown is high.

**Table S1.** Log2 fold change of the significantly differentially Expressed genes identified from different training datasets.

|  | Tissue datasets |  |  | Blood datasets |  |
| --- | --- | --- | --- | --- | --- |
| Gene Symbols | Set 1 | Set 2 | Set 3 | Set 4 | Set 5 |
| DNASE1L3 | NA | -0.0102308 | -0.9566372 | -1.6733822 | -1.7714724 |
| LRRN3 | -1.2863319 | NA | -0.8128004 | -1.4166422 | -1.5215287 |
| SATB1 | -0.6442892 | NA | -1.2565995 | -1.1935436 | -0.9054894 |
| PTGDS | NA | -0.0572214 | -1.5519188 | NA | -2.5739691 |
| EBI3 | NA | -0.0062389 | NA | -1.4540712 | -2.302169 |
| GZMK | NA | -0.1240268 | NA | -1.4072116 | -2.2390033 |
| CTSW | -0.4297419 | -0.0473606 | -1.930786 | NA | -2.2255588 |
| FCMR | -0.9812385 | -0.1511 | NA | -0.7727684 | -1.7854954 |
| CD79A | -0.9141881 | NA | -1.082996 | NA | -1.5609809 |
| GDF10 | NA | -0.0063355 | -1.4747534 | NA | -1.5383126 |
| CD22 | -0.8527311 | -0.0147575 | -1.2656263 | NA | -1.4138414 |
| CD27 | -0.5652415 | -0.1470219 | NA | -0.6619055 | -1.4120213 |
| IL12RB2 | -0.3853798 | NA | NA | -0.9444184 | -1.3933073 |
| CD160 | NA | -0.0872521 | NA | -1.2649517 | -1.3814509 |
| COCH | NA | -0.010724 | -1.5067794 | NA | -1.3124335 |
| NELL2 | -0.9708002 | -0.0632795 | -0.8513571 | NA | -1.2611376 |
| SLAMF1 | -0.4658339 | -0.0380011 | -0.8521605 | NA | -1.1877711 |
| HLA-DPB1 | NA | -0.0203238 | NA | -0.8826801 | -1.1652361 |
| CD3E | -0.4338554 | -0.1082692 | -0.7280421 | NA | -1.1291937 |
| NLGN4X | NA | -0.0069318 | -1.5524494 | NA | -1.1164799 |
| DNAJB9 | NA | -0.0167893 | NA | -0.7593761 | -1.0819126 |
| IL2RB | -0.7635839 | -0.1381649 | -0.7403784 | NA | -1.0255142 |
| CRY2 | -0.2696785 | NA | -1.2989376 | NA | -0.973885 |
| PARM1 | -0.4026864 | -0.0082114 | NA | -1.4185854 | -0.9172337 |
| ACACB | -0.2130128 | NA | -0.8688955 | NA | -0.8474979 |
| NRCAM | -0.4756645 | NA | -0.7238321 | NA | -0.7926464 |
| SPOCK2 | -0.4748107 | -0.0992499 | -0.7014596 | NA | -0.7785083 |
| EIF2AK3 | -0.4004008 | -0.0219009 | NA | -0.5540014 | -0.7118028 |
| SMARCA2 | -0.2730799 | -0.0460831 | NA | -0.8401909 | -0.6641317 |
| PRNP | NA | -0.0618262 | -0.5526453 | NA | -0.4949372 |
| SARAF | NA | -0.1104759 | NA | -0.4872521 | -0.4830139 |
| ASIP | NA | -0.0089826 | -2.1325008 | -2.2481357 | NA |
| CD226 | NA | -0.0155533 | -0.7242504 | -1.8787667 | NA |
| FZD3 | -0.264414 | -0.0079939 | -1.3388057 | -1.5341284 | NA |
| FAM171A1 | NA | -0.0244005 | -0.9119432 | -0.9800952 | NA |
| RNPEP | NA | 0.06499014 | 0.95398998 | NA | 0.41768265 |
| PLOD1 | NA | 0.09547286 | NA | 0.92561755 | 0.50155107 |
| SLC10A3 | NA | 0.01763271 | NA | 0.74658099 | 0.50682203 |
| CTSD | NA | 0.04746329 | 1.2754561 | NA | 0.76021333 |
| FZD2 | NA | 0.02164895 | 1.44884731 | NA | 0.81139532 |
| F11R | NA | 0.01195715 | NA | 0.95747149 | 0.83218316 |
| MET | NA | 0.01887576 | NA | 0.89777193 | 0.85066331 |
| PCDH7 | 0.22264695 | NA | NA | 1.36629866 | 1.01153058 |
| HTATIP2 | NA | 0.02361367 | 0.70979447 | NA | 1.02897248 |
| ECM1 | NA | 0.01714136 | NA | 1.17384734 | 1.18387031 |
| NDNF | 0.33029215 | NA | NA | 1.48913214 | 1.25959925 |
| TINAGL1 | NA | 0.00767782 | NA | 1.35358756 | 1.3607891 |
| EFNA4 | NA | 0.01499515 | 1.54675037 | NA | 1.53163682 |
| TMEM158 | NA | 0.11370092 | NA | 1.93498007 | 1.63762385 |
| DMBT1 | 0.20609413 | NA | NA | 2.37426481 | 1.68706202 |
| CA9 | NA | 0.00649849 | NA | 2.23365804 | 1.699295 |
| DUOX1 | NA | 0.00887676 | NA | 2.44828895 | 2.01800441 |
| KLK7 | NA | 0.00652333 | NA | 4.27690315 | 2.61510498 |
| TFF3 | NA | 0.02998763 | NA | 1.36976068 | 3.02308923 |
| MUC4 | NA | 0.01660057 | NA | 4.34028652 | 4.77504924 |
| CEACAM6 | 0.68734494 | NA | NA | 1.84579246 | 5.37084254 |
| MICB | NA | 0.0454302 | 1.12708641 | 0.61441869 | NA |
| GGH | 0.40428577 | NA | 1.16431707 | 0.64016283 | NA |
| IL1R2 | NA | 0.02805492 | 1.96252861 | 1.19676844 | NA |
| CTSA | NA | 0.06486968 | 1.12882668 | 0.569448 | 0.56228617 |
| ITGB5 | 0.40532311 | NA | 0.56996513 | 0.86378621 | 0.89056218 |
| CD55 | 0.43910958 | NA | 1.68442144 | 1.38634247 | 1.18032619 |
| FAT1 | NA | 0.00801813 | 1.05838548 | 1.09973351 | 1.34356191 |
| SLC6A8 | NA | 0.07658205 | 0.88715672 | 2.45194083 | 1.69464796 |
| SPINT2 | 0.21089938 | NA | 1.52394086 | 1.45628448 | 1.81526649 |
| F12 | NA | 0.01065864 | 1.57281184 | 2.89125329 | 2.09305047 |
| PI3 | NA | 0.13098904 | 1.54565261 | 3.08918788 | 2.97440508 |
| LAMC2 | NA | 0.00581006 | 1.15880058 | 2.4392472 | 3.28863854 |
| ADAM9 | 0.65589477 | 0.01143644 | 1.21182415 | NA | 1.03384287 |
| PLBD1 | 0.98046509 | 0.10857842 | 1.51463411 | NA | 1.38322127 |
| CTSE | 0.55488335 | 0.01164965 | NA | 2.39668584 | 4.75791587 |
| FZD5 | 0.17583608 | 0.00912522 | 0.88362041 | 1.10056425 | 0.74346978 |
| CDCP1 | 0.17986381 | 0.01064018 | 1.35564396 | 1.10288462 | 1.45556502 |
| IFI27 | 0.49426769 | 0.11556995 | 2.84247197 | 2.16446631 | 1.84500054 |

**Table S2:** Direction of differentially upregulated genes validated via boxplot analysis. Upregulated are shown with green background and ones with opposite direction are colored black.

|  | Tissue datasets |  |  | Blood datasets |  |
| --- | --- | --- | --- | --- | --- |
|  | Set 1 | Set 2 | Set 3 | Set 4 | Set 5 |
| RNPEP | Up | Up | Up | Up | Up |
| PLOD1 | Up | Up | Up | Up | Up |
| CTSD | Up | Up | Up | Up | Up |
| FZD2 | Up | Up | Up | Up | Up |
| F11R | Up | Up | Up | Up | Up |
| PCDH7 | Up | Up | Up | Up | Up |
| HTATIP2 | Up | Up | Up | Up | Up |
| EFNA4 | Up | Up | Up | Up | Up |
| DUOX1 | Up | Up | Up | Up | Up |
| KLK7 | Up | Up | Up | Up | Up |
| MUC4 | Up | Up | Up | Up | Up |
| CEACAM6 | Up | Up | Up | Up | Up |
| GGH | Up | Up | Up | Up | Up |
| IL1R2 | Up | Up | Up | Up | Up |
| CTSA | Up | Up | Up | Up | Up |
| ITGB5 | Up | Up | Up | Up | Up |
| FAT1 | Up | Up | Up | Up | Up |
| SLC6A8 | Up | Up | Up | Up | Up |
| SPINT2 | Up | Up | Up | Up | Up |
| F12 | Up | Up | Up | Up | Up |
| PI3 | Up | Up | Up | Up | Up |
| LAMC2 | Up | Up | Up | Up | Up |
| ADAM9 | Up | Up | Up | Up | Up |
| PLBD1 | Up | Up | Up | Up | Up |
| CTSE | Up | Up | Up | Up | Up |
| FZD5 | Up | Up | Up | Up | Up |
| IFI27 | Up | Up | Up | Up | Up |
| SLC10A3 | Up | Up | Up |  | Up |
| TMEM158 | Up | Up | Up |  | Up |
| MICB | Up | Up | Up |  | Up |
| CD55 | Up | Up | Up | Up |  |
| CDCP1 | Up | Up | Up | Up |  |
| MET | Up | Up |  | Up | Up |
| NDNF | Up | Up |  | Up | Up |
| TINAGL1 | Up | Up |  | Up | Up |
| DMBT1 | Up | Up |  | Up | Up |
| CA9 | Up | Up |  | Up | Up |
| TFF3 | Up | Up |  | Up | Up |
| ECM1 | Up | Up |  | Up |  |

**Table S3:** Comparative performance of 9-gene PDAC Classifier with different previously established biomarkers in training, test and validation datasets. Sets with green background are datasets derived from blood. All mustard colored cells have AUC > 0.80 whereas light blue cells indicate low specificity or sensitivity despite of high AUC. For black shaded cells all the genes corresponding to the mentioned studies cannot be identified.

|  |  | Current |  |  |  | Bhasin et al |  |  |  | Balasenthil et al |  |  |  | Kisiel et al |  |  |  | Immunovia |  |  |  |
| --- | --- | --- | --- | --- | --- | --- | --- | --- | --- | --- | --- | --- | --- | --- | --- | --- | --- | --- | --- | --- | --- |
|  |  | Acc | Sens | Spec | AUC | Acc | Sens | Spec | AUC | Acc | Sens | Spec | AUC | Acc | Sens | Spec | AUC | Acc | Sens | Spec | AUC |
| TRAINING | Set 1 | 1 | 1 | 1 | 1 | 0.91 | 1 | 0.83 | 1 | 0.71 | 0.6 | 0.83 | 0.76 | 0.63 | 0.6 | 0.67 | 0.73 | 0.8 | 0.6 | 1 | 1 |
|  | Set 2 | 1 | 1 | 1 | 1 | 1 | 1 | 1 | 1 | 0.5 | 1 | 0 | 0.57 | 0.5 | 1 | 0 | 0.07 | 1 | 1 | 1 | 1 |
|  | Set 3 | 0.87 | 0.83 | 0.9 | 0.89 | 0.95 | 1 | 0.9 | 0.98 | 0.47 | 0.83 | 0.1 | 0.16 | 0.2 | 0.41 | 0 | 0.15 | 0.78 | 0.67 | 0.9 | 0.92 |
|  | Set 4 | 0.82 | 0.93 | 0.71 | 0.93 | 0.49 | 0.97 | 0 | 0.12 | 0.5 | 1 | 0 | 0.01 | 0.5 | 1 | 0 | 0.35 | 0.72 | 0.88 | 0.57 | 0.77 |
|  | Set 5 | 0.86 | 0.89 | 0.89 | 0.97 | 0.5 | 0.45 | 0.56 | 0.53 | 0.78 | 0.78 | 0.78 | 0.81 | 0.47 | 0.44 | 0.5 | 0.49 | 0.59 | 0.56 | 0.62 | 0.64 |
| TEST | Set 6 | 1 | 1 | 1 | 1 | 1 | 1 | 1 | 1 | 0.66 | 0.83 | 0.5 | 0.64 | 0.91 | 1 | 0.83 | 1 | 1 | 1 | 1 | 1 |
|  | Set 7 | 0.92 | 0.9 | 0.93 | 0.94 | 1 | 1 | 1 | 1 | 0.6 | 0.9 | 0.25 | 0.7 | 0.88 | 0.95 | 0.8 | 0.86 | 1 | 1 | 1 | 0.99 |
|  | Set 8 | 1 | 1 | 1 | 1 | 1 | 1 | 1 | 1 | 1 | 1 | 1 | 1 | 1 | 1 | 1 | 1 | 0.91 | 1 | 0.83 | 1 |
|  | Set 9 | 0.95 | 0.91 | 1 | 1 | 1 | 1 | 1 | 1 | 0.9 | 0.75 | 1 | 0.85 | 0.75 | 0.37 | 1 | 0.93 | 0.9 | 0.75 | 1 | 1 |
|  | Set 10 | 0.96 | 0.93 | 1 | 0.94 | 0.93 | 0.8 | 1 | 0.87 | 0.75 | 0.2 | 1 | 0.71 | 0.83 | 0.46 | 1 | 0.89 | 1 | 1 | 1 | 0.92 |
|  | Set 11 | 0.73 | 0.75 | 0.71 | 0.8 | 0.47 | 0 | 0.93 | 0.22 | 0.71 | 0.58 | 0.85 | 0.65 | 0.51 | 0.16 | 0.85 | 0.23 | 0.29 | 0.09 | 0.5 | 0.18 |
| VALIDATION | V1 | 0.79 | 0.76 | 0.83 | 0.83 | 0.84 | 0.86 | 0.82 | 0.92 | 0.6 | 0.21 | 0.95 | 0.72 | 0.76 | 0.62 | 0.88 | 0.7 | 0.89 | 0.77 | 1 | 0.94 |
|  | V2 | 0.98 | 0.97 | 1 | 1 | 0.98 | 0.95 | 1 | 0.99 | 1 | 1 | 1 | 1 | 0.9 | 0.85 | 1 | 0.9 | 1 | 1 | 1 | 1 |
|  | V3 | 0.94 | 1 | 0.89 | 0.98 | 0.98 | 0.88 | 1 | 0.99 | 0.96 | 0.77 | 1 | 0.99 | 0.96 | 0.77 | 1 | 0.96 | 0.94 | 0.66 | 1 | 0.96 |
|  | V4 | 0.95 | 1 | 0.91 | 0.99 | 0.99 | 0.91 | 1 | 1 |  |  |  |  | 0.99 | 0.91 | 1 | 0.99 |  |  |  |  |
|  | V5 | 0.83 | 0.84 | 0.82 | 0.89 |  |  |  |  | 0.67 | 0.96 | 0.24 | 0.82 |  |  |  |  |  |  |  |  |

